# Subthalamic nucleus encoding steers adaptive therapies for gait in Parkinson’s disease

**DOI:** 10.1101/2025.08.20.25333478

**Authors:** Stefano Scafa, Valeria de Seta, Ruijia Wang, Paula Sánchez López, Camille Varescon, Icare Sakr, Nadia Bérard, Lea Bole-Feysot, Céline Deschenaux, Ian Enderli, Andrea Sánchez López, Yohann Thenaisie, Morgane Burri, Frédéric Merlos, Vanessa Fleury, Ettore Accolla, Benoit Wicki, Cécile Hübsch, Mayte Castro Jiménez, Julien F. Bally, Alessandro Puiatti, Kyuhwa Lee, Henri Lorach, Antoine Collomb-Clerc, Gregoire Courtine, Jocelyne Bloch, Eduardo M. Moraud

**Author notes:** contributed equally to this work (Stefano Scafa, Valeria de Seta, Ruijia Wang, Jocelyne Bloch, Eduardo M. Moraud).

## Abstract

Parkinson’s disease leads to a spectrum of cardinal motor symptoms and locomotor deficits that vary in severity with the nature of daily activities and the fluctuating physiology of patients. Many of these deficits remain inadequately addressed by existing therapies that use continuous, activity-agnostic parameters. Instead, adaptive therapies embedding activity-specific parameters have the potential to better address the entire range of symptoms. Here, we expose physiological principles that enable real-time decoding of ongoing locomotor activities across motor fluctuations from the neural dynamics of the subthalamic nucleus. This decoding steered activity-dependent adaptations of deep brain stimulation therapies that improved both cardinal motor symptoms and locomotor deficits across activities of daily living. Our decoding framework provides a blueprint for next-generation neuromodulation therapies that continuously adapt parameters to the behavioral context and fluctuating physiology of each patient.

**One Sentence Summary:** Neural decoders that leverage the physiological principles of activity-dependent encoding in the subthalamic nucleus support the implementation of adaptive deep brain stimulation therapies that alleviate locomotor deficits in people with Parkinson’s disease.

## INTRODUCTION

Parkinson’s disease (PD) leads to a spectrum of locomotor deficits, including heterogeneous gait abnormalities, balance disturbances, and freezing of gait^1,2^. While some of these symptoms improve with dopamine replacement pharmacotherapy (L-DOPA) and deep brain stimulation (DBS) of the subthalamic nucleus (STN), many locomotor deficits remain refractory to these treatments^3,4^. In late-stage PD, up to 90% of individuals experience therapy-resistant locomotor impairments that markedly increase the risk of falls, lead to a loss of independence, and reduce quality of life^5,6^. Addressing these deficits remains a priority in the treatment of PD^7^.

Conventional DBS therapies employ stimulation parameters that are primarily optimized to alleviate rigidity, bradykinesia, and tremor^8,9^. These parameters are often suboptimal to alleviate locomotor deficits, and may even aggravate symptoms such as postural instability and freezing of gait^3^. Clinical observations suggest that when DBS parameters are tuned to target locomotor deficits instead of cardinal motor signs, they can mitigate these symptoms^9–11^. These observations suggest that optimal DBS therapies must incorporate multiple configurations of parameters tailored to both cardinal motor symptoms and locomotor deficits across daily activities. Moreover, dopamine-related fluctuations and context-dependent physical and cognitive demands can dramatically influence the nature and severity of locomotor deficits^12,13^. We therefore anticipate that optimal DBS therapies must not only incorporate additional parameters addressing gait and balance deficits, but that these parameters must also be adapted in real time based on the ongoing activity and fluctuating state of each patient.

Developing such therapies requires the identification of physiological principles that reveal patient-specific encoding of locomotor activities across motor fluctuations, to enable activity-dependent DBS adaptations in real time. The STN is a central hub within the neuronal architecture that encodes motor function and dysfunction, both while sitting and across mobility activities^14–16^. We previously demonstrated that locomotor activities and gait deficits can be decoded from local field potentials (LFPs) recorded via DBS electrodes implanted in the STN of people with PD^17^. Moreover, implantable neuro-stimulation platforms endowed with sensing functionalities now enable chronic recordings of STN LFPs to deliver adaptive therapies throughout daily life^18–20^. Therefore, the STN is not only a physiologically-relevant but also a technologically-accessible target to extract principles for steering adaptive neuromodulation therapies. Indeed, the identification of principles encoding bradykinesia and rigidity within STN dynamics^21–23^ already enabled adaptive DBS strategies operating in closed-loop that outperform conventional DBS protocols targeting these symptoms^24–27^.

We reasoned that the STN is also a natural and practical target to identify physiological principles that can steer adaptive neuromodulation therapies for gait and balance^28–31^. However, these principles remain unknown. Here, we aimed to uncover these principles. We show that the distinct muscle activation demands underlying daily mobility activities are encoded within STN dynamics. Machine learning strategies informed by these principles guided the implementation of activity-dependent DBS therapies, personalized to each patient, that ameliorated both cardinal motor symptoms and locomotor deficits in real time.

## RESULTS

### Multimodal platform to dissect activity-dependent STN dynamics

We aimed to uncover the physiological principles by which STN dynamics encode patient-specific locomotor activities underlying daily mobility, and how this encoding is altered by conventional therapies, including L-DOPA and STN DBS.

We anticipated that uncovering these principles would require mapping electrophysiological signals recorded from the STN onto precise behavioral measurements collected across a range of locomotor activities and therapeutic conditions. To enable this mapping, we established a wireless platform integrating an implantable pulse generator with sensing capabilities for real-time recording of STN LFPs (*26*), and high-resolution tracking of full-body kinematics and lower-limb muscle activity during unconstrained mobility (**Fig. 1a** and **Supplementary Video 1**).

**Figure 1.**
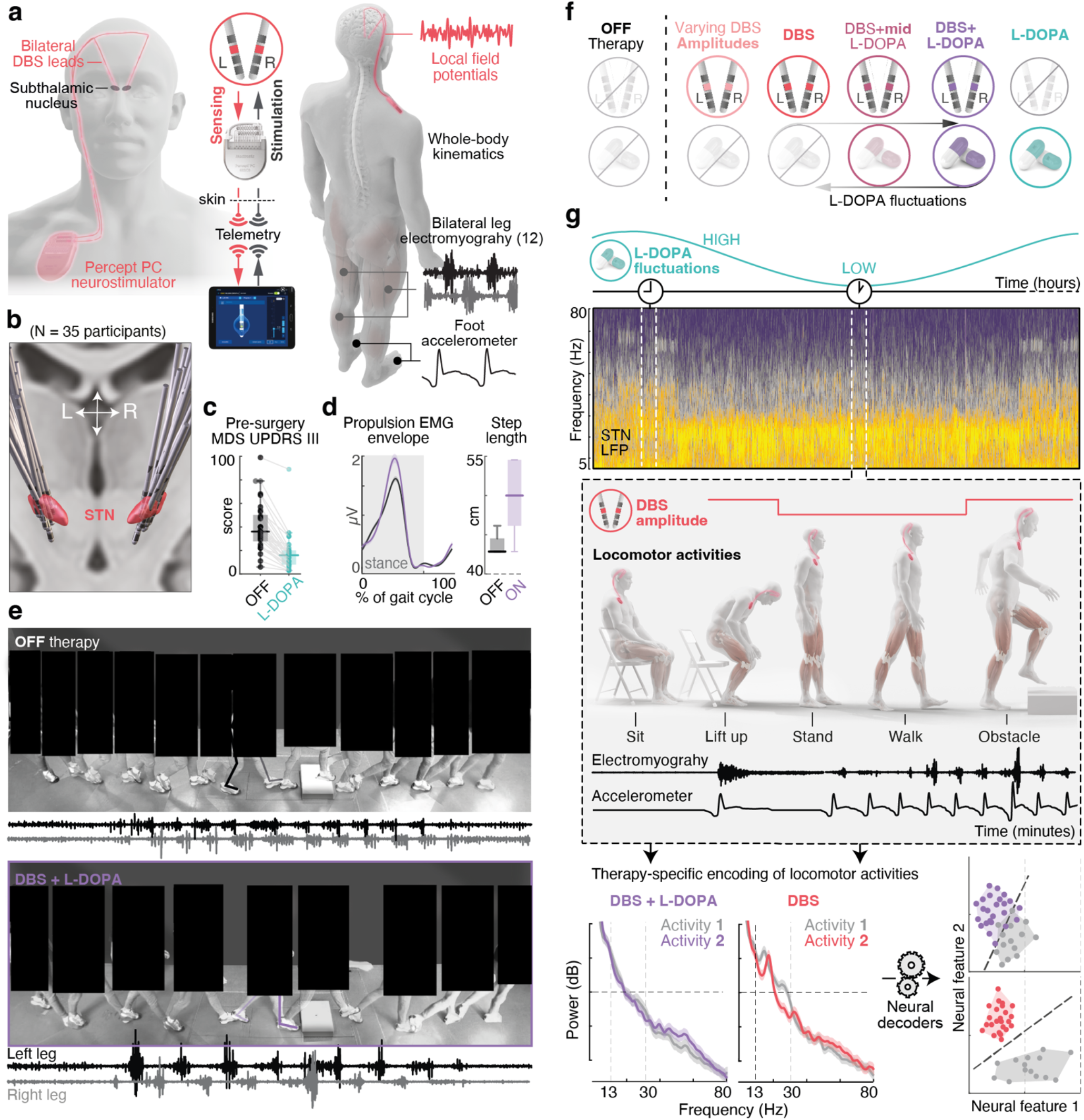
Experimental setup to dissect activity-dependent STN dynamics across therapeutic conditions. **(a)** Wireless monitoring platform enabling continuous recording of STN local field potentials, bilateral leg muscle activity, and whole-body kinematics during unconstrained mobility in PD participants implanted with a last-generation DBS neurostimulator with sensing capabilities. **(b)** Anatomical reconstruction of lead placements across the 35 participants included in the study. **(c)** Clinical profiles of included participants. Boxplots show pre-surgery clinical scores (MDS-UPDRS III) measured in the OFF-therapy condition (no L-DOPA for >12 h), and their improvement in response to L-DOPA. **(d)** Representative metrics quantifying gait improvements induced by the standard-of-care combination of STN DBS and L-DOPA (ON-therapy condition) versus the OFF-therapy condition after DBS implantation in one illustrative participant. Traces represent the mean ± sem. of EMG envelopes from the lateral gastrocnemius muscle, normalized to gait cycle duration. Boxplots show changes in step length. **(e)** Chronophotography of an obstacle avoidance sequence in the OFF therapy condition (*top*) and under standard-of-care STN DBS and L-DOPA (*bottom*), aligned to knee extensor muscle activity (Vastus Medialis, VM) for the right and left legs. **(f)** Therapeutic conditions studied include distinct combinations of STN DBS and L-DOPA, designed to isolate their individual contributions as well as their interaction over time. **(g)** Experimental protocol to dissect the physiological and therapeutic components required to implement activity-dependent DBS therapies for real-world applications. The protocol involves mapping STN neural dynamics to locomotor activities across L-DOPA fluctuations and changes in DBS amplitude. To capture these dynamics, assessments are performed at selected time windows throughout the L-DOPA cycle. Locomotor performance is evaluated under different DBS amplitudes. Neural data collected during these periods are used to train decoders capable of automatically discriminating locomotor states across varying dopaminergic and stimulation conditions.

We enrolled 35 individuals with advanced PD who exhibited motor fluctuations (MDS UPDRS III median score = 36 OFF L-DOPA, median score improvement with L-DOPA = 20, **Fig. 1b-c** and **Extended Data Table 1**). Kinematic quantifications confirmed the prevalence of locomotor impairments in this cohort (**Fig. 1d,e**). All participants underwent bilateral implantation of DBS leads in the STN. Post-operative reconstructions confirmed accurate lead placement (**Fig. 1b** and **Extended Data Fig. 1**).

For each participant, we conducted electrophysiological assessments to define low-beta, high-beta, and gamma frequency bands using parametric fitting methods^1^ (**Extended Data Fig. 2**). We also confirmed that the standard-of-care combination of L-DOPA and STN DBS led to improvements in gait quality (**Fig. 1d,e**).

To dissect the physiological and therapeutic components necessary to guide the implementation of activity-dependent therapies for gait, we mapped neural dynamics to mobility activities across conditions encountered in daily life: (i) the standard-of-care combination of L-DOPA and STN DBS; (ii) L-DOPA and STN DBS administered independently; (iii) varying dosages of both therapies; and (iv) the OFF-therapy condition as a baseline for comparison (**Fig. 1f,g** and **Extended Table 2**).

To address the complexity of interactions between experimental paradigms and therapeutic conditions, participants were divided into four groups, each assigned to specific protocols (see **Methods** and **Extended Data Table 2**).

### STN dynamics encode locomotor activities in the absence of therapy

Daily mobility encompasses a diversity of locomotor activities, such as standing, walking, turning or navigating environmental constraints—each demanding distinct and often asymmetric activation of bilateral leg muscles (**Fig. 2a**). We previously showed that the STN encodes the timing and vigor of muscle activation during isolated leg movements^17^. We thus hypothesized that each of these locomotor activities must also be encoded by specific signatures within STN dynamics.

**Figure 2.**
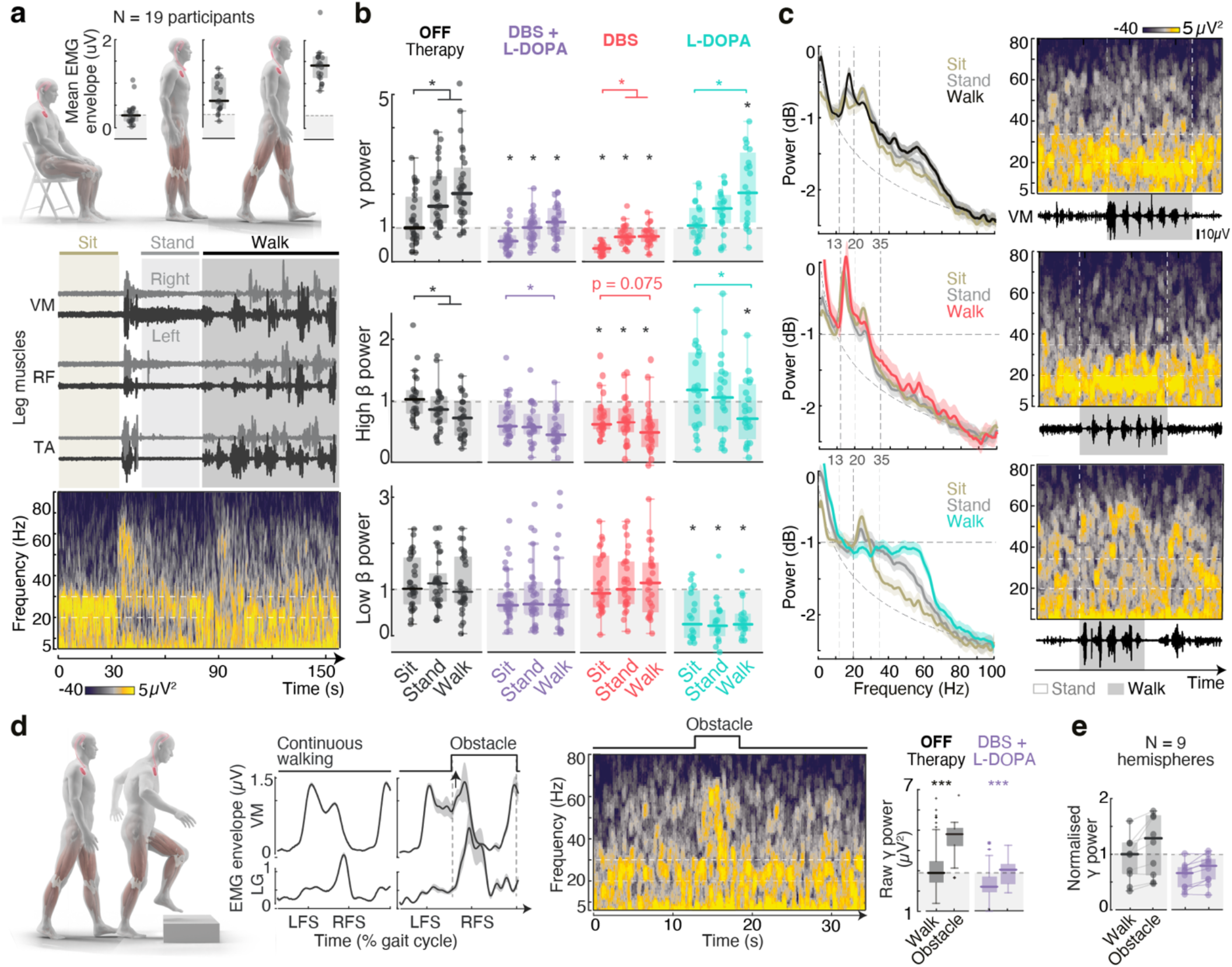
Therapy-dependent encoding of locomotor activities within STN dynamics. **(a)** Illustrative example of EMG modulations for three bilateral leg muscles (Vastus Medialis VM, Rectus Femoris RF, and Tibialis Anterior TA) aligned to the time-resolved spectrogram of STN LFP during transitions from sitting to standing to walking. Boxplots represent the mean EMG activation (computed from bilateral VM envelopes), showing a progressive increase in motor demands across locomotor activities. **(b)** Boxplots showing band-power modulations across locomotor activities in the low-beta (mean frequency range: 12.9–19.5 Hz), high-beta (21.6–31.6 Hz), and gamma (37.7–68.8 Hz) bands under each therapeutic condition. Power values are normalized to the group median at rest in the OFF-therapy condition. Dots represent mean power values for each individual hemisphere included in the analysis (N = 32 hemispheres OFF therapy; N = 32 ON DBS+L-DOPA; N = 27 ON DBS; N = 17 ON L-DOPA). Colored stars indicate significant differences between locomotor activities (sitting, standing, and walking) within the same therapeutic condition. Black stars above individual boxplots denote statistically significant differences relative to the OFF-therapy condition for the same subset of hemispheres. **(c)** (*Left*) Power spectral density (PSD) of STN LFPs across different locomotor activities in the OFF-therapy condition, under STN DBS alone, and under L-DOPA alone, shown for an illustrative participant (PD139). (*Right*) Corresponding time-resolved spectrogram aligned to leg EMG activity (Vastus Medialis, VM) during a sequence of interleaved walking and standing.**(d)** *(Left)* Average EMG envelope profiles (mean ± sem) of two leg muscles (knee extensor VM, ankle extensor LG) normalized to the gait-cycle duration, shown for continuous walking (left) and obstacle avoidance (right). Stepping over the obstacle elicits increased leg muscle activation to stabilize the stance leg and propel the body while lifting the contralateral leg (LFS: Left footstrike, RFS: right footstrike). *(Right)* Representative example of spectral modulation during a walking sequence that includes an obstacle avoidance step, and boxplots of modulations in raw gamma power for the same participant. **(e)** Cross-participant analysis of normalized gamma-band power across the five participants who successfully completed the task. * p<0.05, *** p<0.001, Bonferroni correction for multiple comparisons.

To test this hypothesis, we mapped modulations in STN LFPs to leg muscle activation during sitting, standing, walking, and obstacle avoidance. We first conducted this mapping in the absence of therapy. As expected, these activities elicited a graded increase in the level of bilateral leg muscle activation that reflected progressive adaptations in the range and complexity of leg movements (**Fig. 2a**). This activity-dependent increase in muscle activation coincided with a progressive increase in gamma-band power, accompanied by a gradual decrease in high-beta power (black boxplots and traces in **Fig. 2b,c**). These reproducible patterns contrasted with heterogeneous modulations in low-beta power, which varied across participants. Indeed, scrutiny of low-beta patterns revealed participant-specific increases, decreases, or no consistent changes across the 19 individuals (**Fig. 2b** and **Extended Data Fig. 2, 3a,b)**. Similar gamma band modulations were observed during the phase of increased leg muscle activation required for obstacle avoidance (**Fig. 2d,e**).

These results showed that, in the absence of therapy, STN dynamics encode the various locomotor activities involved in daily mobility through specific spectral modulations.

### Standard-of-care therapy alters activity-dependent STN dynamics

L-DOPA and STN DBS are routinely combined to manage the symptomatology of people with PD. However, this standard-of-care therapy also modulates STN dynamics, potentially altering the encoding of locomotor activities and limiting the suitability of these signals for steering adaptive therapies. Since our goal is to develop adaptive DBS protocols under the influence of both treatments, we next asked whether the delivery of STN DBS and L-DOPA with standard-of-care parameters alters activity-dependent STN dynamics.

We first confirmed that L-DOPA and STN DBS impro-ved mobility in our cohort of participants. Quantifications of kinematics and muscle activity revealed clear improvements in the quality and vigor of gait patterns across the studied locomotor activities (**Fig. 1d**). We next examined concomitant changes in STN dynamics. Despite a pronounced reduction in baseline (sitting) power levels, the gradual modulations in gamma and high-beta power during transitions from sitting to standing to walking were preserved. However, the amplitude of these activity-dependent modulations was markedly less pronounced than in the absence of therapy (purple boxplots in **Fig. 2b** and **Extended Data Fig. 2 and 3a,b**). Inspection of low-beta power confirmed the persistent absence of reproducible modulation patterns across participants (**Extended Data Fig. 2, 3a**).

While these results confirmed that the encoding of locomotor activities within STN dynamics is preserved under L-DOPA and STN DBS, they also revealed that the combination of these therapies markedly weakens the strength of this encoding.

### Therapy-specific alterations in activity-dependent STN dynamics

While both L-DOPA and STN DBS alleviate locomotor deficits, clinical observations indicate that they exert distinct effects on specific gait and balance impairments. We reasoned that these differences must reflect distinct therapy-specific alterations in activity-dependent STN dynamics^32,33^. Dissecting these alterations is essential to inform targeted adaptations of DBS parameters in patients treated with both therapies

We found that L-DOPA versus STN DBS altered activity-dependent STN dynamics in strikingly opposite directions. L-DOPA abolished low-beta power modulations across all studied locomotor activities, while concurrently amplifying activity-dependent modulations in high-beta and gamma power (turquoi--se boxplots and traces **Fig. 2b,c, Extended Data Fig. 3c,d)**. These changes altered the contribution of these bands to the encoding of locomotor activities. In contrast, STN DBS reduced power levels in the high-beta and gamma bands across all activities, thus weakening their role in this encoding (red boxplots and traces in **Fig. 2b,c** and **Extended Data Fig. 3c,d** and **Supplementary Video 1**).

These results emphasize that therapy-specific alterations in activity-dependent STN dynamics must be incorporated in the design of algorithms that steer adaptive therapies.

### Impact of L-DOPA pharmacokinetics on activity-dependent STN dynamics

L-DOPA pharmacokinetics drive variations in brain dopamine levels, which induce motor fluctuations throughout the day. We anticipated that these same fluctuations must also impose time-dependent alterations in the encoding of locomotor activities within STN dynamics.

To dissect these alterations, we evaluated changes in STN dynamics during sitting, standing and walking every 15 minutes over the course of one hour following the administration of L-DOPA. STN DBS remained activated throughout these evaluations. As expected, L-DOPA mediated progressive improvements in gait parameters over the studied period (**Fig. 3a**). These behavioral improvements were associated with a gradual shift in the contribution of the spectral bands that encode locomotor activities (**Fig. 3b)**. Concretely, L-DOPA mediated a gradual reduction in low-beta and high-beta power, together with a progressive increase in gamma power, both of which correlated with gait quality (**Fig. 3c,d**).

**Figure 3.**
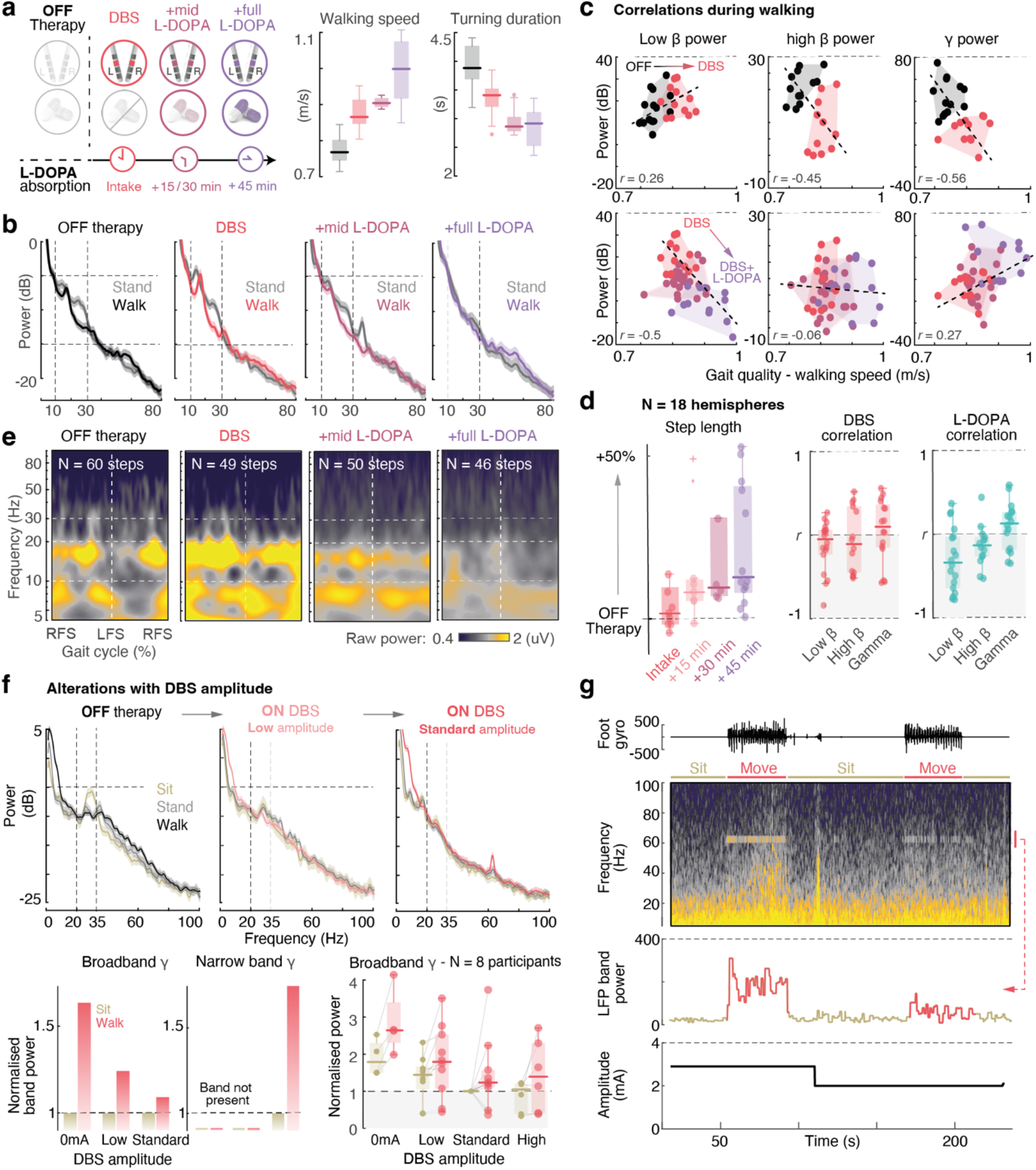
Progressive modulation of activity-dependent STN dynamics with L-DOPA pharmacokinetics and DBS amplitude. **(a)** Experimental protocol to assess the evolving interaction between STN DBS and L-DOPA over time, including gait evaluations at 15, 30, and 45 minutes following L-DOPA intake. Boxplots show key gait quality parameters across conditions for participant PD101, illustrating progressive improvements with therapies. **(b)** Illustrative examples of power spectral densities showing activity-dependent modulations between standing and walking, and their progressive alteration with STN DBS and L-DOPA pharmacokinetics in the same participant. Dashed vertical lines indicate the beta frequency band. **(c)** Correlation analyses between average band-power and gait quality (walking speed) for the low-beta, high-beta and gamma bands across therapeutic conditions. The individual effects of STN DBS (top row) and the subsequent effects of L-DOPA absorption alongside STN DBS (bottom row) illustrate distinct behaviors over time. Each dot represents the mean band power per walking sequence (10 sequences performed in each condition). Dotted lines represent linear correlations. **(d)** (*Left*) Boxplots display cross-participant changes in step length for each therapeutic condition (normalized to OFF-therapy). (*Right*) Correlation scores (*r*) between gait quality (walking speed) and band power across all N = 11 participants, measured under STN DBS or throughout L-DOPA absorption. L-DOPA induces strong band-specific modulations in STN LFPs that correlate with gait improvements (primarily negative correlations in the low-beta band and positive correlations in the gamma band), while correlations with STN DBS are variable and highly patient-dependent. **(e)** Time-resolved spectral decomposition averaged across gait-cycles, interpolated between consecutive contralateral footstrikes, for the same participant and under the same therapeutic conditions shown in (b). Dashed horizontal lines highlight low and high beta bands (see also **Extended Data Fig. 3**). RFS = Right footstrike, LFS = Left footstrike. **(f) (***Top*) Representative examples of power spectral density from one participant (PD130), showing activity-dependent modulations across three DBS amplitudes: the standard-of-care amplitude, half that amplitude, and 0 mA. Frequency-band encoding changes progressively with stimulation. Broadband gamma modulation gradually decreases and is replaced by a narrowband gamma peak that also encodes locomotor activities. (*Bottom*) Bar plots quantify gamma power modulation across all three conditions, normalized to the resting (sitting) state in the same participant. Box plots show broadband gamma modulations across the eight participants who underwent these assessments. Not all participants could be recorded at 0 mA, and not all hemispheres displayed gamma-band modulation (N = 4 hemispheres for 0 mA, N = 9 for low amplitude, N = 10 for the standard-of-care amplitude, and N = 6 for high amplitude). **(g)** Illustrative example of narrowband gamma modulation with movement and DBS amplitude (participant P3, DBS frequency: 125 Hz). Spectrogram (*top*) shows pronounced narrowband gamma activity during movement, which diminishes as DBS amplitude is reduced over time (*bottom*); STN LFP band power in the 60–65 Hz range as computed in real time by the implanted DBS platform (*middle*). This narrowband gamma modulation was not observed across all participants.

Time–frequency analyses averaged over each gait cycle showed that the gradual reduction in low-beta power during L-DOPA absorption was due to weaker bursts of synchrony during the propulsion phase of walking^17^ (**Fig. 3e** and **Extended Data Fig. 3a)**. In contrast, STN DBS variably increased or decreased the amplitude of these low-beta bursts across participants (**Fig. 3e**).

These results exposed the time-dependent impact of L-DOPA on STN dynamics, revealing the need for adaptive therapies to maintain stability despite continuously evolving encoding of locomotor activities.

### Impact of DBS amplitude on activity-dependent STN dynamics

Activity-dependent DBS strategies aim to adjust stimulation parameters in real time based on ongoing locomotor activities. Clinical experience has shown that stimulation amplitude is a sensitive parameter to modulate different motor symptoms and is routinely adjusted as a first-line approach^9^. This makes DBS amplitude a logical first target for real-time control. However, since changes in DBS amplitude can also influence STN dynamics, we next assessed their impact on the encoding of locomotor activities.

In the tested cohort, increasing DBS amplitude led to progressive reductions in gamma power, both at baseline and during activity-related modulations. In several participants, this suppression was accompanied by the emergence of a narrow gamma band at half the stimulation frequency, which also exhibited modulations with locomotor activities (**Fig. 3f,g** and **Extended Data Fig. 3e)**.

These observations indicate that adjustments in amplitude can shift the frequency band through which locomotor activities are encoded. Adaptive DBS therapies must therefore account for these gradual, amplitude-dependent spectral transitions when designing robust control algorithms.

### Therapy-specific decoders predict locomotor activities

We next asked whether these physiological principles could support the development of machine learning strategies to guide activity-dependent DBS therapies. Given the opposing impact of L-DOPA versus STN DBS on activity-dependent STN dynamics, we reasoned that that these therapies may hinder the stability of neural decoders across therapeutic conditions. To address this question, we asked whether a neural decoder trained to discriminate locomotor activities in the absence of therapy remains accurate when standard-of-care L-DOPA and STN DBS is administered.

We developed a machine learning pipeline based on personalized Random Forest classification algorithms^34^ that were trained to decode three key classes of locomotor activities: sitting, standing and walking (**Fig. 4a**). In the absence of therapies, the decoders classified the three locomotor activities with high accuracy during cross-validation assessments (**Fig. 4b-d**). However, the performance of these decoders declined dramatically when L-DOPA and STN DBS were administered (**Fig. 4b** and **Extended Data Fig. 4a,b**). This decline reflected the expected shift in activity-dependent spectral features when delivering the standard-of-care therapy, which led to overlaps between the spectral signatures of different classes (**Fig. 4e** and **Extended Data Fig. 4c-e**). To compensate for these overlaps, we trained separate decoders using STN LFPs collected under the standard-of-care combination of L-DOPA and STN DBS, or under STN DBS alone (**Extended Data Fig. 4j)**. These therapy-specific decoders restored accurate classification of locomotor activities under their respective therapeutic condition, but failed to generalize across conditions (**Fig. 4b** and **Extended Data Fig. 4a,b**). Feature contribution analyses revealed that each decoder relied on distinct spectral bands **(Fig. 4e** and **Extended Data Fig. 4c-e)**.

**Figure 4.**
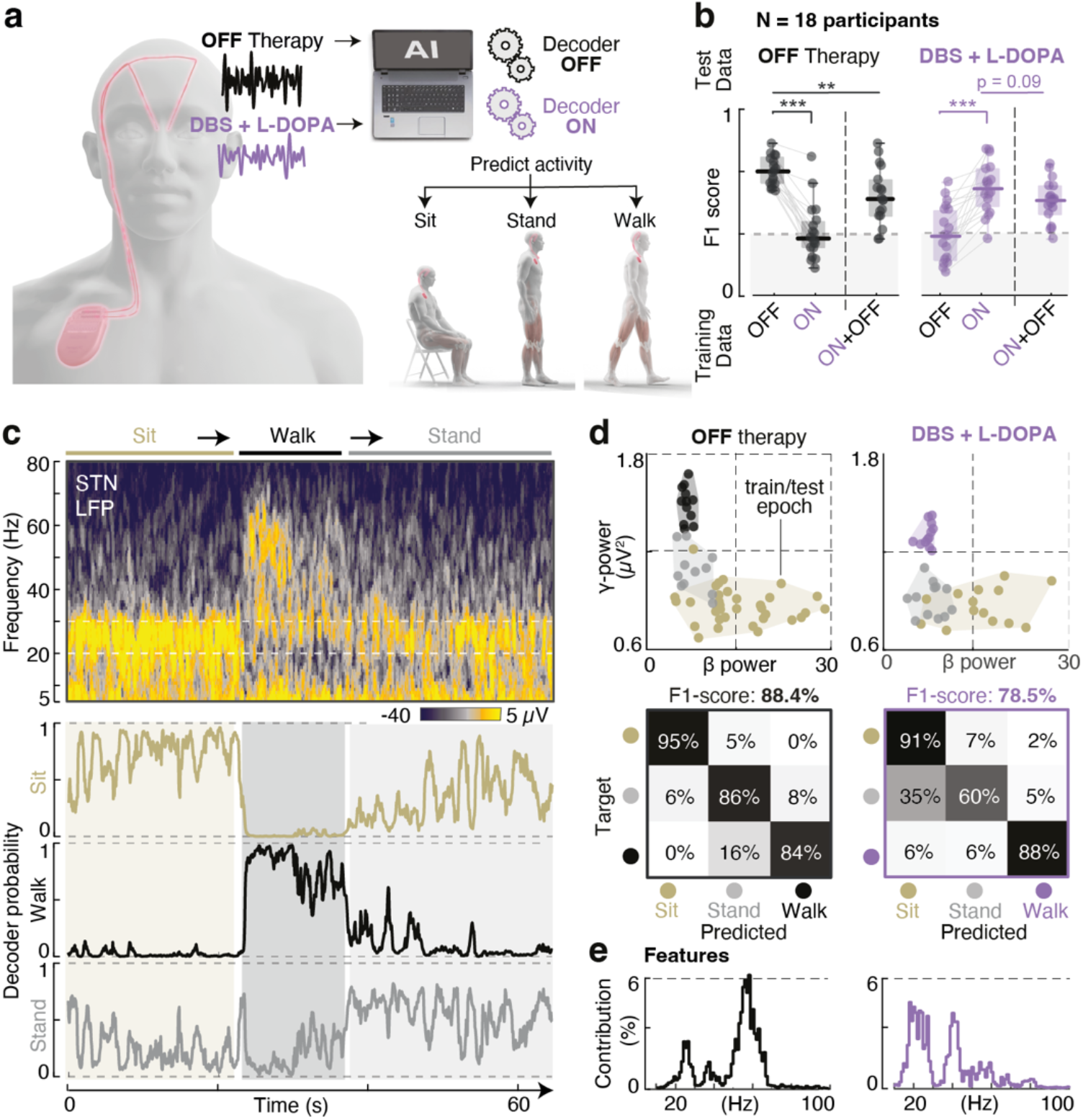
Robust neural decoding of locomotor activities across therapeutic conditions requires therapy-specific decoders. **(a)** Computational setup to decode locomotor activities (sit, stand, walk) from STN LFP acquired in the OFF-therapy condition and under the standard-of-care combination of STN DBS and L-DOPA (ON-therapy condition). Two decoders were trained independently. Their performance was then compared across both conditions. **(b)** Boxplots of average decoding performance (F1-score) across all 18 participants. Three decoders are compared: two decoders trained on data from either the OFF-therapy condition (decoder OFF) or the standard-of-care combination of L-DOPA and STN DBS (decoder ON), and a third decoder trained on data from both conditions together (decoder ON+OFF). Each decoder was then tested under each therapeutic condition. **(c)** Illustrative example of neural decoding during a sit–stand–walk transition in the OFF-therapy condition. Raw spectrogram and decoder probability traces are shown for a decoder trained and tested on data recorded under the same therapeutic condition (participant PD121). **(d)** *(Left)* Scatter plot of average band-power distributions for the beta and gamma bands shows the separability between locomotor states (same participant) in the OFF-therapy condition. Each dot represents an epoch used for training or testing the decoder. Confusion matrix of a therapy-specific decoder confirms correct classification of all three states (F1-score = 88.4%). *(Right)* Same representation for the DBS + L-DOPA condition, illustrating the shift in the distributions of each class in the 2D space, and confusion matrix for a therapy-specific decoder trained in this condition (F1-score = 78.5%). **(e)** Spectral feature traces contributing to each therapy-specific decoder, illustrating the distinct frequency bands that most effectively support decoding under each therapeutic condition (same participant). ** p<0.01, *** p<0.001 one-way ANOVA with Bonferroni correction.

We finally asked whether a single decoder trained on STN LFPs collected across all therapeutic conditions –without therapy and under standard-of-care therapy— could generalize across conditions. This unified decoder failed to classify locomotor activities accurately, due to overlapping activity-specific spectral features that a single decoder could not disentangle (**Fig. 4b** and **Extended Data Fig. 4f,g**). Similarly, training across participants failed to generalize to individual patients, under-scoring the need for personalized decoders (**Extended Data Fig. 4h,i**).

These results demonstrate that locomotor activities can be predicted from STN dynamics with accuracy, but that therapy-specific shifts in activity-dependent spectral encoding prevent a single decoder from generalizing across therapeutic conditions.

### Modular decoding preserves performance across therapeutic conditions

Our results indicate that therapy-specific decoders are necessary to maximize classification accuracy. However, fluctuations in L-DOPA levels and changes in DBS amplitude impose time-varying alterations in STN dynamics. We therefore anticipate that continuous decoding across therapeutic conditions will require combining multiple decoders, each selected according to the ongoing therapeutic state. To test this possibility, we implemented a modular framework that integrates two therapy-specific decoders and a classifier that dynamically switches between them.

We first tested whether this modular framework maintains accurate decoding of locomotor activities despite L-DOPA-related fluctuations (**Fig. 5a**). We trained two decoders, one under L-DOPA and STN DBS, and one under STN DBS alone (**Fig. 5b** and **Extended Data Fig. 6a**). We then optimized the classifier to estimate the probability of being under the influence of L-DOPA (**Fig. 5c**). Because L-DOPA fluctuations unfold over slower timescales than locomotor-related modulations, we tuned the classifier’s hyper-parameters to filter out fast, movement-related dynamics while preserving slower, L-DOPA-induced changes. This tuning was guided by a genetic algorithm (GA) that identified the optimal hyper-parameter configuration (**Extended Data Fig. 7**). On all 9 participants tested, the classifier predicted the medication state, despite individual differences in absorption times (**Fig. 5d,e**). In turn, the modular framework enabled accurate decoding of locomotor activities despite the progressive shift in activity-dependent spectral encoding due to the gradual absorption of L-DOPA over the duration of the recordings (**Fig. 5d,e, Extended Data Fig. 6a** and **Supplementary video 2**).

**Figure 5.**
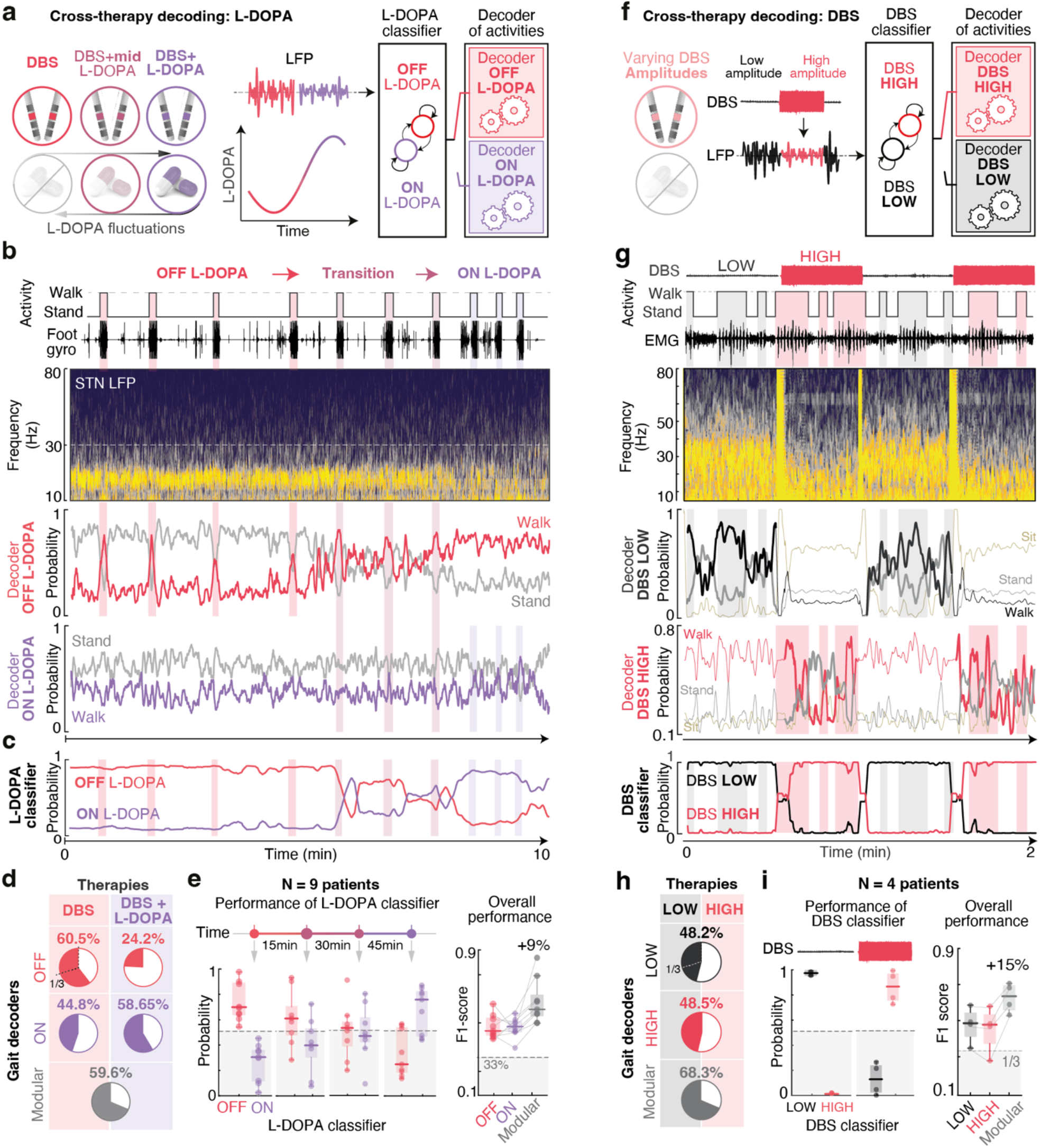
A modular decoding framework maintains prediction accuracy across therapeutic conditions. **(a)** A modular decoding framework predicts locomotor activities across L-DOPA pharmacokinetics. Two parallel decoders are trained independently, one under DBS alone (decoder OFF L-DOPA) and another trained under combined DBS + L-DOPA (decoder ON L-DOPA). An intermediate layer (L-DOPA-classifier) selects the appropriate decoder based on L-DOPA-related features of the incoming STN LFP signal. The hyperparameters of the classifier are optimized to filter out fast, locomotor-related modulations while preserving slower, L-DOPA-induced changes in STN LFPs; this tuning is performed using a genetic algorithm that identifies the parameter configuration best suited for discriminating ON and OFF L-DOPA states (**Extended Data Fig. 7**). **(b)** Representative example of raw spectrogram (top) and probability traces from each decoder (bottom) during ten back-and-forth walking sequences over a 10-minute task, initiated 15 minutes after L-DOPA intake (participant PD104). As L-DOPA is progressively absorbed, STN dynamics and decoder performance evolve. **(c)** Probability traces of the L-DOPA classifier over the course of the same recording. **(d)** Average decoder performance (F1-score) under DBS alone or under DBS + L-DOPA conditions, highlighting the limited cross-condition generalizability of each decoder (decoder OFF L-DOPA mean F1-score = 42.35%, decoder ON L-DOPA mean F1-score = 51.7%). The modular framework markedly improves overall decoding accuracy across conditions. **(e)** (*Left*) Boxplots show the performance of the L-DOPA classifier at 15, 30, and 45 minutes post-L-DOPA intake across 9 participants, illustrating the classifier’s ability to capture the gradual absorption of L-DOPA. (*Right*) Improvement in decoding performance across participants when using the modular framework compared to either decoder alone. **(f)** A modular decoding framework predicts locomotor activities across two DBS amplitudes. Two parallel decoders are trained independently: one at the standard-of-care stimulation amplitude (decoder DBS HIGH) and one at 0 mA (decoder DBS LOW). A DBS classifier selects the appropriate decoder based on stimulation-related features in STN LFPs. The hyperparameters of the classifier are optimized using a genetic algorithm to detect rapid, stimulation-induced changes while ignoring slower, activity-related modulations. **(g)** Illustrative example of DBS profile (*top*), knee extensor EMG aligned to raw STN LFP spectrogram (*middle*), and decoder probability traces (*bottom*) during repeated walking sequences (participant PD130). DBS amplitude was manually toggled between low and high values independently of task execution. **(h)** Pie charts show average decoder performance (F1-score) across the full trial. The modular framework improves decoding accuracy. **(i)** Boxplots showing classification accuracy and decoding performance (F1-score and median probability values) across all 4 participants tested. * *p*<0.05 one-way ANOVA with Bonferroni correction.

We then asked whether this modular framework is also effective to maintain accurate decoding of locomotor activities during adaptations of DBS amplitudes (**Fig. 5f** and **Extended Data Fig. 6b**). For this, we trained the classifier to detect the amplitude of STN DBS, and implemented two therapy-specific decoders to predict locomotor activities under high DBS amplitude (standard-of-care) or low DBS amplitude (0 mA) (**Fig. 5g**). Despite the expected alteration of activity-dependent STN dynamics with changing DBS amplitude, the modular framework maintained high decoding accuracy (**Fig. 5h,i** and **Extended Data Fig. 6a**).

We finally assessed the robustness of the modular decoding framework in real-life settings (**Extended Data Fig. 8**). Locomotor activities were monitored using sensorized shoes that continuously computed spatiotemporal gait parameters as participants ambulated for extended periods across indoor and outdoor environments (**Extended Data Fig. 8a**). These ecological assessments involved frequent transitions between sitting, standing, and walking in crowded corridors, elevators, and while navigating obstacles. Despite these variable environmental conditions, the modular framework maintained accurate decoding of locomotor activities across therapeutic conditions (**Extended Data Fig. 8b,c**).

This decoding framework provides a blueprint to leverage the physiological principles underlying therapy-specific encoding of locomotor activities, and to steer adaptive therapies that adjust stimulation parameters to the behavioral context and fluctuating physiology of each patient.

### Activity-dependent DBS therapies alleviate locomotor deficits

Finally, we sought to establish proof-of-concept for the potential of this decoding framework to steer activity-dependent DBS therapies targeting both cardinal motor symptoms and locomotor deficits.

The physiological principles underlying the encoding of locomotor activities and their modulation by therapies revealed that robust discrimination of activities across therapeutic conditions requires considering the full spectrum of frequency bands, beyond the beta band typically accessible in commercial devices. Furthermore, therapies must incorporate versatility in the control of DBS parameters to target either cardinal motor symptoms or locomotor deficits. We translated these requirements into an investigational neurostimulation platform that supports up- and down-regulation of stimulation amplitude guided by continuous monitoring of a spectral feature selected across the entire STN LFP frequency spectrum (**Fig. 6a,b** and **Extended Data Fig. 9a,b**).

**Figure 6.**
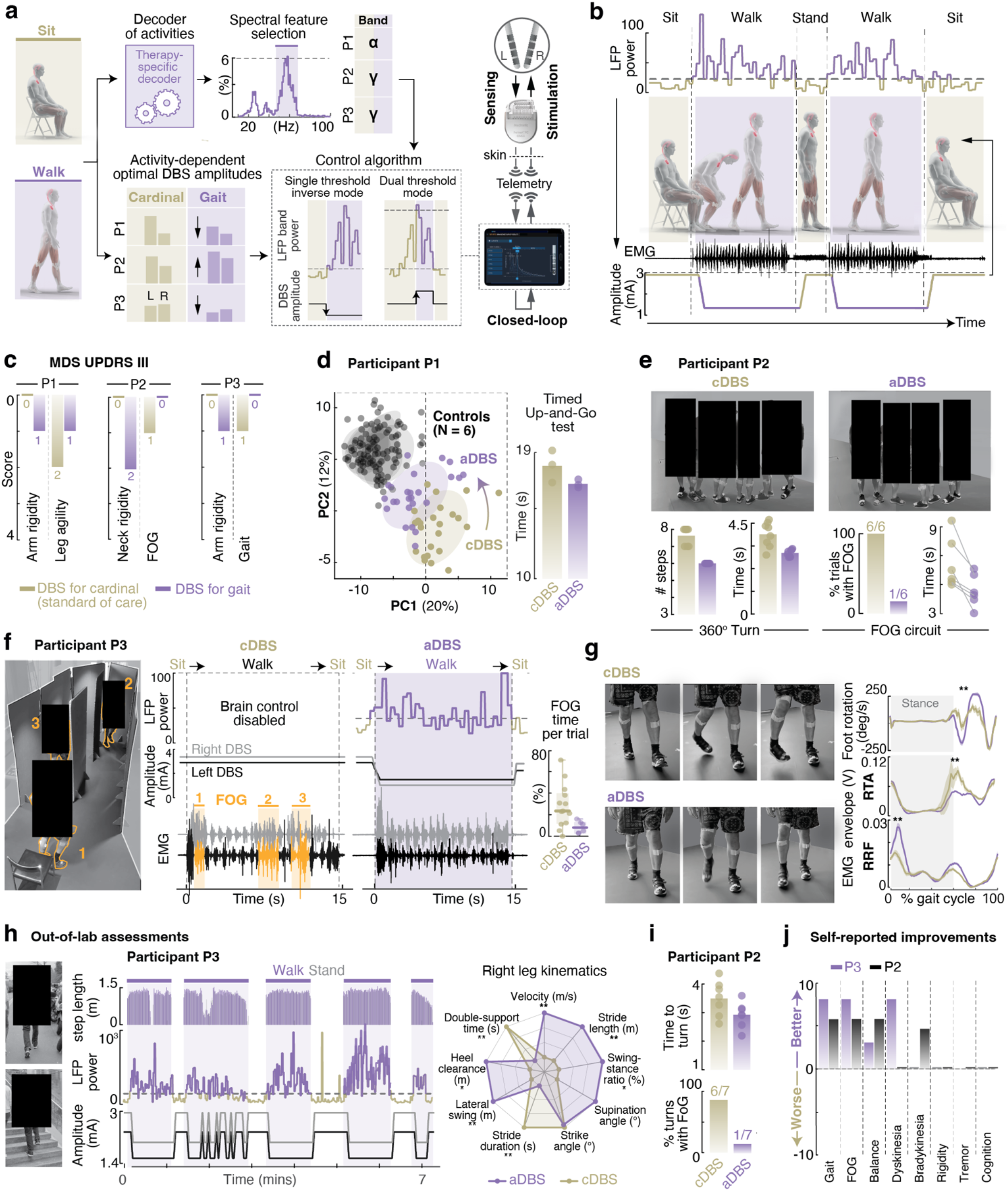
Activity-dependent DBS therapies alleviate locomotor deficits and cardinal motor symtoms. **(a)** Experimental setup for implementing activity-dependent DBS protocols using an investigational DBS platform with real-time closed-loop capabilities. Therapy-specific decoders identify the most robust frequency band for discriminating locomotor activities in each participant (i.e., alpha band for participant P1, and gamma band for participants P2 and P3). In parallel, optimal DBS amplitudes for alleviating the most bothersome cardinal motor symptoms and locomotor deficits are determined individually and used to guide algorithm selection. These protocols are implemented in an investigational DBS platform with closed-loop capabilities, including up- or down-regulation of stimulation amplitude in real time. **(b)** Representative example of activity-dependent DBS adaptations during a task involving alternations between sitting, standing, and walking. Power in the selected LFP band is monitored in real time and used to trigger adaptations between DBS amplitudes optimized for cardinal motor symptoms versus locomotor impairments. **(c)** MDS-UPDRS III scores for the most bothersome cardinal versus locomotor deficits under each set of DBS amplitudes. Selected items included upper-limb and neck rigidity (as participants presented with akinetic-rigid syndromes) compared to gait disturbances, freezing of gait, and lower-limb agility. **(d)** (*Left*) Principal component analysis (PCA) of gait quality for participant P1 under standard-of-care continuous DBS (cDBS) and activity-dependent adaptive DBS (aDBS), compared to healthy controls. Each dot represents a single gait cycle, parameterized by 77 features and projected onto a two-dimensional space defined by the first two principal components (PC1 and PC2). Feature contributions to each component are shown in **Extended Data Fig. 11a**. (*Right*) Time to complete three repetitions of the Timed-Up-and-Go (TUG) task under each stimulation condition. **(e)** *(Top)* Chronophotography of a 360° turning task performed by participant P2 under standard-of-care DBS (cDBS) and activity-dependent DBS (aDBS). (*Bottom*) Barplots quantify improvements in turning and reduced freezing of gait with activity-dependent DBS (same participant), both in the context of a 360° turning task and a customized FOG circuit (see also **Extended Data Fig. 11c**). **(f)** Chronophotography of a freezing-of-gait circuit task, which involves navigating a narrow figure-eight corridor, in participant P3. In the low L-DOPA condition, this participant exhibited multiple FOG episodes, including during transitions from sit-to-stand and at recurrent locations along the circuit. Activity-dependent reductions in DBS amplitude mitigated the occurrence of FOG and improved task performance, as shown by representative raw EMG traces under both conditions. Boxplot quantifies improvements across multiple repetitions of the same task under each condition (N=13 under cDBS, and N=6 under aDBS). **(g)** Participant P3 exhibited disabling right foot and leg dyskinesia in the high L-DOPA condition that severely disrupted locomotion. Activity-dependent reductions in DBS amplitude mitigated these impairments, improving leg muscle activation profiles and foot kinematics (median ± sem). PCA analysis for the same task is shown in **Extended Data Fig. 11b. (h)** (*Top*) Out-of-laboratory assessments confirmed the robustness and sustained efficacy of activity-dependent DBS in improving gait patterns in participant P3. Panels show aligned step-length trajectories. (*Bottom*) LFP band power used to trigger DBS amplitude adaptations and corresponding stimulation adjustments. Temporal parameters of the aDBS algorithm (average duration, blanking period, and onset delay) are tuned to maximize the stability of adaptations. Radard plots show kinematic gait features extracted from sensorized shoes (NUSHU, Magnes AG, Switzerland), which further supported improvements in gait quality under activity-dependent DBS. **(i)** In participant P2, out-of-laboratory testing confirmed that activity-dependent DBS preserves efficacy for reducing FOG and improving turning (see also **Extended Data Fig. 11d**). **(j)** Self-reported assessments conducted during out-of-laboratory sessions confirmed perceived improve-ments in overall motor function, primarily driven by gait-related benefits without worsening of cardinal motor symptoms. * *p*<0.05, ** <*p*<0.01 t-test with Bonferroni correction.

We conducted a feasibility clinical trial in three participants with advanced PD and severe motor fluctuations, whose gait impairments persisted despite optimized DBS settings (clinicaltrial.gov, NCT06791902, **Extended Data Table 3** and **Extended Data Fig. 10a,b**). Despite clear motor improvements following STN DBS lead implantation, all participants experienced a progressive worsening of cardinal motor symptoms over time, which was managed through iterative adjustments of stimulation parameters. However, these adjustments were suboptimal to mitigate locomotor deficits and, in some cases, even aggravated these deficits. We therefore investigated whether adjusting DBS amplitudes differentially improved locomotor deficits versus cardinal symptoms. For each participant, we characterized their most debilitating gait impairments and identified optimized DBS amplitudes to alleviate these deficits versus cardinal symptoms (**Fig. 6a,c** and **Extended Data Tables 3 and 4**).

Participant P1 presented with pronounced upper-limb rigidity that was managed with high DBS amplitudes, but these parameters blocked the right leg during walking. Reducing DBS amplitude mitigated these interferences (**Fig. 6c,d)**. Participant P2 displayed severe freezing of gait when low in L-DOPA, especially when turning, passing through doors, and avoiding obstacles. Increasing DBS amplitude mitigated these difficulties, but exacerbated neck rigidity and impaired speech (**Fig. 6c,e)**. Participant P3 exhibited prominent L-DOPA-related dyskinesias that affected the right lower-limb and disrupted walking. Reducing DBS amplitude immediately alleviated these deficits, but worsened upper-limb rigidity. Participant P3 also experienced freezing of gait when low in L-DOPA. During experimental sessions, FOG improved by reducing DBS to very low amplitudes (**Fig. 6c,f,g**).

Having identified activity-specific DBS parameters, we next aimed to determine patient-specific neural signatures capable of capturing ongoing activities across varying therapeutic conditions. Since our investigational neurostimulation platform only allowed monitoring one pre-defined spectral feature per hemisphere, we used our decoding framework to compute the most discriminative frequency to predict locomotor activities (**Extended Data Fig. 10c-e**). We computed this optimal feature for each hemisphere, separately for high and low L-DOPA conditions, but across DBS amplitudes to ensure stability during real-time control of stimulation amplitude. Optimal spectral features typically differed between high and low L-DOPA conditions and across hemispheres (**Extended Data Fig. 10c-e** and **Extended Data Table 4**).

We finally embedded these personalized parameters into adaptive DBS protocols that adjusted stimulation amplitude in real time based on activity-dependent modulations of the optimal spectral feature. We tested the preliminary safety and efficacy of these protocols in well-controlled laboratory settings (**Fig. 6d-g** and **Extended Data Fig. 11a-d**), and then in out-of-laboratory conditions (**Fig. 6h-j** and **Extended Data Fig. 11e**). Activity-dependent adaptations in DBS parameters not only addressed cardinal symptoms when patients were sitting or standing, but also mitigated locomotor deficits during walking (**Fig. 6d-g** and **Supplementary video 3**). All participants exhibited improvements in their most debilitating locomotor deficits. Participant P1 showed improved leg agility during walking (**Fig. 6d** and **Extended Data Fig. 11a)**, P2 experienced reduced freezing of gait (**Fig. 6e** and **Extended Data Fig. 11c)**, and P3 exhibited alleviation of leg dyskinesias and freezing of gait (**Fig. 6f,g** and **Extended Data Fig. 11b**), with no worsening of cardinal symptoms at rest in any participant. Overall, these improvements ameliorated leg muscle activation patterns, symmetry, and gait fluidity. Questionnaires assessing parti-cipants’ subjective experience during out-of-laboratory assessments confirmed the superiority of adaptive DBS over conventional DBS protocols (**Fig. 6j**).

## DISCUSSION

We exposed physiological principles by which the STN encodes locomotor activities across motor fluctuations in people with PD. This understanding guided the development of a decoding framework capable of predicting locomotor activities in real time from STN LFPs, regardless of L-DOPA fluctuations and changes in DBS parameters. We leveraged this framework to implement personalized, activity-dependent DBS protocols on an investigational neurostimulation platform with closed-loop capabilities. In a proof-of-concept clinical study in three individuals with severe gait and balance impairments, activitydependent DBS improved both locomotor deficits and cardinal motor symptoms in real-world settings.

Conventional DBS therapies are primarily optimized to alleviate rigidity, bradykinesia, or tremor. However, optimal management of both cardinal motor symptoms and locomotor deficits requires stimulation configurations tailored to each set of impairments. Since the severity of these deficits vary with the ongoing daily activities and fluctuating physiology, effective adaptations of stimulation are contingent on real-time detection of ongoing mobility activities. Our analyses in a cohort of 35 PD patients revealed that the distinct muscle activation demands underlying these activities are encoded in STN dynamics. These findings confirm that the STN encodes vigor during whole-body movements, consistent with prior reports on upper-limb movements^35,36^ and isolated leg muscle activation^17^, and extend this principle to complex locomotor activities and therapeutic conditions. Indeed, we found that locomotor encoding was retained under both L-DOPA and STN DBS, despite profound and opposing therapy-specific alterations that primarily affected low-beta and gamma frequency bands^32,33,37^. These alterations hindered neural decoder performance across therapeutic conditions.

We addressed this challenge with a modular decoding framework composed of therapy-specific decoders operating in parallel. This modular architecture decomposed a complex, multi-objective control problem into smaller, therapy-specific sub-problems, which achieved reliable prediction of locomotor activities even in the presence of L-DOPA motor fluctuations and DBS adjustments. To disentangle neural modulations associated with locomotor activities from therapy-induced alterations in the same STN LFP signals, each module was tuned to operate over a distinct timescale, corresponding to the temporal dynamics of locomotion, L-DOPA fluctuations, or DBS-induced changes.

This modular architecture is well suited for medical applications that impose strict technical and clinical constraints^38^. Algorithms embedded in implantable devices must be computationally efficient, easy to tune across individuals, and trainable with limited amounts of data. Ideally, decoder parameters should also remain interpretable by healthcare professionals, empowering them to personalize therapies independently. In our implementation, each therapy-specific decoder required only a small number of free parameters and could be trained within minutes using a few sequences of locomotor activities. All other hyperparameters were optimized offline using a genetic algorithm to ensure accuracy and transferability across patients and therapies. This modular design also permits the seamless integration of additional inputs as new modules, such as electrocorticography signals^26,39,40^ or data from wearable sensors^41^, thereby maximizing flexibility and scalability.

We leveraged the modular decoding framework to guide the implementation of activity-dependent adaptive DBS strategies targeting both cardinal motor symptoms and locomotor deficits in real time. We obtained proof of concept for the efficacy of these strategies in a clinical study involving three participants with persistent cardinal symptoms and locomotor deficits. The identification of patient-specific spectral features that capture ongoing locomotor activities despite the confounding effects of L-DOPA and STN DBS was essential for delivering robust adaptations in stimulation parameters^42^. Our results showed that this identification required exploiting the rich spectral encoding of locomotor activities under each therapy, which spans multiple—and often overlapping—frequency bands across the entire STN LFP spectrum. This included frequencies beyond the beta-band typically accessible in commercial devices^43^.

We implemented these strategies on an investigational neurostimulation platform that not only harnessed the full spectral content of STN LFPs for advanced monitoring and control, but also enabled band-specific up- or down-regulation of stimulation amplitude. These functionalities were pivotal to meet the specific requirements of locomotor impairments. While the three tested participants exhibited heterogeneous clinical profiles and substantial motor fluctuations, activity-dependent adaptations of DBS parameters ameliorated the full spectrum of deficits. These results demonstrate the clinical feasibility of activity-dependent DBS therapies to address gait impairments in addition to cardinal motor symptoms. However, reliance on a single frequency band limited robustness for long-term use in real-world conditions. While our modular decoding framework helped address this limitation, expanding feedback to multiple frequency bands or incorporating additional inputs beyond STN LFPs will further enhance control stability^27^. Moreover, although adaptations in stimulation amplitude mitigated gait impairments in our three participants, future platforms may additionally need to support adjustments in stimulation frequency and contact configuration to enable activity-dependent delivery of lower frequencies via ventral contacts that better target gait-related pathways^44^.

Our modular decoding framework offers a blueprint for next-generation neuromodulation therapies that dynamically adapt stimulation parameters to the behavioral context, mobility demands, and fluctuating physiology of each patient^45,46^. Realizing the full potential of activity-dependent adaptive DBS will require dedicated hardware and large-scale clinical trials to demonstrate long-term safety and efficacy in real-life settings.

## Data Availability

All data associated with this study are present in the paper or the Extended Data Materials. Anonymized data that support the findings of this study and custom code used will be made available upon reasonable request to the corresponding author

## MATERIALS AND METHODS

### Study design

The objective of this study was to uncover the physiological principles by which the subthalamic nucleus encodes daily mobility activities across motor fluctuations in people with Parkinson’s disease, and to leverage this understanding to develop activity-dependent DBS therapies capable of alleviating both cardinal motor symptoms and locomotor deficits in real time.

The study was conducted in two parts. First, we characterized how locomotor activities are represented within STN dynamics and how these neural patterns are altered by L-DOPA and STN DBS. Assessments in 35 participants informed the development of neural decoders to guide the implementation of adaptive DBS strategies. Second, we conducted a feasibility clinical trial (NCT06791902) using an investigational neurostimulation platform (Medtronic, USA) to evaluate the safety and preliminary efficacy of activity-dependent DBS in three individuals with PD.

### Part 1 | Characterisation of physiological principles and development of neural decoders

41 participants with Parkinson’s disease were recruited for this part of the study. All participants were implanted with bilateral deep brain stimulation directional leads (Sensight, Medtronic, USA) and a Percept PC implantable neurostimulator (Medtronic, USA) in the right pectoral area. All participants were recorded in the seven days that followed their DBS surgery using the sensing capabilities offered by the neurostimulator. Participants recorded in the OFF-medication condition were withdrawn from their usual dopaminergic medication the night before the recordings (>12h). For recordings ON DBS or ON L-DOPA, stimulation amplitudes and medication dosages were kept as defined optimal by the Neurologist at the time of the recording (**Extended Data Table 1)**.

#### Patient exclusion

Two participants were unable to complete the required set of motor tasks due to poor physical condition and were not recorded. Among the remaining 39 participants that were recorded, three participants had to be excluded from analyses because all LFP channels exhibited severe ECG-related artifacts that rendered signals unusable. One additional participant had to be excluded since the “cycling” mode of his neurostimulator was activated, which made it impossible to compare different therapeutic conditions in a clean manner. Overall, 35 participants were retained for the analyses.

#### Participation to motor tasks

Not all 35 patients participated in all motor tasks. Participants were grouped based on the therapeutic conditions and motor tasks in which they were assessed (**Extended Data Table 2**):

- Group A included 11 participants who performed continuous walking tasks under five therapeutic conditions: (i) OFF therapy, (ii) ON DBS only, and (iii) ON DBS + L-DOPA at three time points following medication intake (15, 30, and 45 minutes).
- Group B involved 13 participants who performed an obstacle-avoidance walking task under two conditions: (i) OFF therapy and (ii) standard-of-care combination of STN DBS + L-DOPA.
- Group C consisted of 8 participants who completed continuous walking tasks in two therapeutic conditions: (i) OFF therapy and (ii) ON L-DOPA only. Two additional participants of group B performed these tasks.
- Group D comprised 3 participants who completed the walking task under varying DBS amplitudes. In addition, the 7 participants from the feasibility clinical study on adaptive DBS also performed this task and were included in the corresponding analyses.

During walking tasks, one participant from Group A was excluded due to incorrect DBS parameter settings during the experimental session. Additionally, four participants did not receive L-DOPA and were therefore only assessed in the corresponding experimental conditions. In Group D, one participant was stimulated at 85 Hz, which prevented assessment of gamma-band modulations across varying DBS amplitudes. Another participant in this group completed experiments at low DBS amplitudes, but not at their standard clinical setting.

As a result, the number of participants retained for each task was as follows: 19 participants were analyzed during walking tasks under standard therapeutic conditions (OFF-therapy and the standard-of-care combination of STN DBS and L-DOPA); 11 participants during walking tasks tracking the time-dependent effects of L-DOPA pharmacokinetics (15, 30, and 45 minutes post-intake); 15 and 10 participants during walking tasks under STN DBS only and L-DOPA only, respectively; and 8 participants under varying DBS amplitudes. Additionally, 4 participants performed walking tasks during manual toggling of DBS amplitude to evaluate the online robustness of neural decoders

All experiments were approved by the Ethical Committee of the Canton de Vaud, Switzerland (ref PB_2017-00064). Informed consent was obtained before each recording. The nature and possible consequences of the study were systematically explained to all participants.

### Part 2 | Feasibility clinical trial to assess activity-dependent adaptive DBS

Seven individuals with advanced PD were screened for enrollment in the study. All screened participants had received bilateral DBS leads in the STN (Sensight, Medtronic, USA) and a Percept PC neuro-stimulator (Medtronic, USA) in the right chest area at least three months prior to the screening. Despite optimized standard therapy, they continued to experience disabling gait impairments.

Each participant underwent an initial session to assess eligibility for inclusion in the trial (see below). Three individuals met all inclusion criteria and proceeded to the full experimental protocol. Demographic and clinical data are detailed in **Extended Data Table 3** and **Extended Data Fig. 10b**. At the time of enrollment, DBS parameters and levodopa regimens had been optimized by each participant’s treating neurologist to target cardinal motor symptoms consistent with akinetic-rigid profiles. The median stimulation amplitude across the six implanted hemispheres was 2.9 mA, consistent with clinical therapeutic ranges.

The study was approved by the Swiss Ethics Committee (Swissethics BASEC 2024-D0066, protocol no. 10001378, approved on 21 November 2024) and the Swiss regulatory agency (Swissmedic SNCTP 000006185, protocol no. 102731544, approved on 23 December 2024) and conducted in accordance with the Declaration of Helsinki. The study is registered on ClinicalTrials.gov (NCT06791902). All participants provided written informed consent prior to recruitment and agreed to the publication of identifiable images and videos.

Additionally, to compare participants’ gait quality under activity-dependent and conventional DBS with physiological gait, kinematic data were collected from six healthy controls (3 male, 3 female) walking in a laboratory setting (research protocol ref: 2017-02112, approved by the Ethical Committee of the Canton de Vaud, Switzerland). All participants signed the informed consent before participation.

### DBS electrode localization

Lead localization was computed using the processing pipeline (Horn & Li et al. 2018) in the Lead-DBS Matlab toolbox (Horn & Kühn. 2017), using pre-operative T1 and T2-weighted MRIs and a post-operative CT-scan as inputs. Briefly, post-operative CT scan was linearly co-registered to pre-operative MRI using advanced normalization tools (ANTs, Avants et al. 2011). Coregistrations were visually verified and manually corrected if needed. A brain shift correction step was applied, as implemented in Lead-DBS. All preoperative volumes were used to estimate a precise multi-spectral normalization to ICBM 2009b NLIN asymmetric space (Fonov et al. 2009) applying the ANTs SyN Diffeomorphic Mapping (Avants et al. 2008) using the preset “effective: low variance default + subcortical refinement”. DBS contacts were automatically pre-reconstructed using the phantom-validated and fully automated PaCER method (Husch et al. 2017). They were all individually verified.

Of all recorded hemispheres, seven were excluded from analysis based on anatomical reconstructions raising concerns that the selected contacts were not fully located within the motor subregion of the STN (**Extended Data Fig. 1**). This anatomical uncertainty was corroborated by the absence of any detectable signal modulation across the entire spectrum of STN LFPs or under different therapeutic conditions throughout the experiments.

### LFP recordings

Local field potentials were recorded using the sensing capabilities of the Percept PC neurostimulator in BrainSense™ Streaming mode (sampling rate: 250 Hz). This mode permits sensing from a single bipolar contact pair per hemisphere. For each participant, the contact pair adjacent to the clinically active stimulation electrode (previously selected by the treating neurologist) was systematically chosen for recordings. In cases where stimulation was delivered through contacts incompatible with sensing (contacts 0 or 3), the nearest available contact pair was used.

Synchronization with external systems was achieved by delivering a brief DBS burst (same stimulation parameters as standard-of-care therapy) at the start and end of each recording. These pulses produced identifiable artifacts in an external EMG sensor placed on the chest, near the implantable pulse generator (IPG), enabling precise alignment across recording modalities.

### Definition of LFP power frequency bands

The identification of power frequency bands was personalized for each patient. Low-beta, high-beta, and gamma bands were identified using an unbiased algorithm that fits the PSD of each patient as a combination of an aperiodic (1/f) and a mix of Gaussian components^1^. The algorithm iteratively optimized the fit and found the most appropriate number of Gaussian components and their parameters (mean and SD) to model the original spectrum with a given accuracy.

Since frequency bands may differ or shift between rest and movement, and specific bands may only emerge during movement (such as broadband gamma), we run the fitting algorithm on the PSD computed on a complete set of recordings, including both rest and movement periods. Identified Gaussian components in the range of 10 to 20 Hz were labeled as “low beta,” those in the range of 20 to 35 Hz were labeled as “high beta”, and those above 35Hz as Gamma.

Of the 35 participants assessed (63 hemispheres retained), a low-beta band was identified in 54 hemispheres (mean frequency range: 12.9 Hz - 19.5 Hz), a high-beta band in 58 (mean range: 21.6 Hz - 31.6 Hz), and a gamma band in 56 (mean range: 37.7 Hz - 68.8 Hz) (**Extended Data Fig. 2**). Subsequent calculations of activity-dependent power modulations in each band included only hemispheres in which the corresponding band was identified.

### Removal of ECG-related artifacts from LFP

For participants whose LFP recordings exhibited ECG-related artifacts (N = 10), a denoising algorithm based on singular value decomposition^2^ was applied to isolate and recover the underlying neural signal.

Artifact timing was first identified from an external EMG signal recorded via a chest-mounted sensor, which also served for synchronization purposes. The EMG signal was z-scored, and R-peaks were detected using the ‘*findpeaks’* function (MATLAB R2024a) with the following criteria: (i) a minimum peak amplitude of 2.5 standard deviations above baseline, and (ii) a minimum inter-peak interval of 500 ms. The timing of the detected R-peaks was then projected onto the synchronized LFP signal to isolate QRS complexes. For each peak, we extracted a 400 ms window (200 ms before and after the R-peak) from the LFP and decomposed it using SVD. The ECG artifact was reconstructed using the two singular components with the largest eigenvalues and subtracted from the original LFP signal. A final offset correction was applied to each segment by minimizing the sum of squared errors between the raw signal and the reconstructed artifact. This procedure was applied to participants whose LFPs exhibited ECG contamination (N = 10).

For the few cases where ECG peaks were not captured by the chest EMG sensor, we used the raw LFP signal itself to identify the timing of ECG-induced artefacts. The process to compute ECG complexes and denoise the signal was identical.

### Locomotor tasks

#### Walking tasks

Patients were instructed to stand for 3 seconds before initiating a sustained bout of walking (in a straight line) at their comfortable speed. When arriving at the end of the bout, patients were instructed to stop and stand for 3 seconds, before doing a U-turn and starting another walking bout.

#### Obstacle task

Patients were asked to walk at their natural speed in a straight line (without any marks on the floor). An obstacle (height 10.5cm, length = 39.5cm) was placed on their path. They were requested to stride over it and continue walking normally until the end of the bout.

### Biomechanical recordings during locomotor tasks

#### Kinematics (in-lab)

High-fidelity recordings were carried out using an optoelectronic motion capture system (Vicon, UK) that measured 3D positions of key body joints (foot, ankle, knee, hip). Kinematic data was complemented with bilateral triaxial inertial measurement unit (IMU) sensors (Delsys, USA) attached to the patients’ shoes, which recorded raw gyroscope signals from the right and left feet (sampling frequency: 148Hz).

#### Kinematics (out-of-lab)

Walking periods and stepping quality were recorded using sensorized shoes (Nushu, Magnes, Swizterland) that are endowed with IMUs in the their insole. Embedded machine-learning algorithms automatically detect gait events, and for each gait cycle, they compute parameters such as step-length, step-height, or maximal foot acceleration.

#### Electromyographic signals

EMG signals were recorded using wireless EMG sensors operating at 1.5kHz (Delsys, USA). Sensors were placed bilaterally, on agonist and antagonist muscles of the ankle joint (TA Tibialis Anterior, MG Medial Gastrocnemius, LG Lateral Gastrocnemius), knee joint (VM Vastus Medialis, ST Semitendinosus) and hip joint (RF Rectus Femoris). An additional EMG sensor was placed on the chest for synchronization purposes. All recordings were synchronized using dedicated trigger signals.

### Identification of gait events during walking tasks & definition of locomotor states

#### Bilateral gait events

We computed periods of walking and turning, along with toe-off and heel-strike events, automatically from gyroscope signals from the right and left feet using a graphical tool (http://github.com/matteonocilli/GAIT_tool)3 consisting of three main sections: segmentation of walking periods, gait-event detection, and manual correction. The algorithm was developed in Matlab® R2022a and allows to extract toe-off (TO) and heel-strike (HS) events from angular velocity signals (mid-lateral axis) recorded from gyroscopes placed on the feet.

#### Gait segmentation

Recognition of uninterrupted movement (walking) periods from rest (sitting or standing) was automatically computed from feet accelerations, using a pattern-matching process trained on (and adapted from) a pool of healthy subjects.

#### Definition of exact gait event times

Within the movement phases identified previously, the detection of swing phases was obtained by identifying the main acceleration peak that happens at the detachment of the foot from the ground. Gait events (TO and HS) correspond to the preceding and following minimum acce-lerations, respectively. To avoid mismatch of false swings, a threshold value was set (Th = 2.5x STD), beyond which the peak could be defined as a swing.

#### Manual correction

Corrections were applied by visually inspecting the signal to remove possible outliers.

#### Identification of turning sequences

We used the low-pass filtered gyroscope signals in the coronal plane (5Hz, 5^th^-order Butter-worth filter). Turning peaks needed to be superior to a threshold (defined as 75% quantile of the datapoints exceeding the mean of the whole recording) and at least 0.2 seconds apart. Turning sequences were pooled together: Peaks separated by less than 1.75 times the mean inter-peak distance across the recording were assumed to belong to the same turning sequence.

#### Locomotor states

Using the aforementioned gait events, we categorized each time-point of the recordings into five discrete locomotor states, labeled as “sitting”, “standing”, “continuous walking”, and “turning”. This state-machine description of gait was further used for developing gait-state decoders and computing state-related analyses of power.

### Analyses of gait quality during locomotor tasks

We defined a “walking sequence” as a walking bout, between each stop and turn, and a “gait cycle” as the interval between consecutive foot strikes of the same leg, representing a complete step.

#### Gait parameters

For each walking sequence, we computed (i) average walking speeds by dividing the walking bout distance by the time taken to complete the sequence (measured in meters per second, m/s) and (ii) and the aveage step length as the bout distance divided by the total number of steps in the sequence (measured in meters, m). We defined turning duration as the time taken to perform each turn between walking sequences.

#### Muscle activation

We first computed EMG envelopes for each individual muscle. Raw EMG signals were band-pass filtered (20-500Hz, zero-lag 4^th^-order Butterworth filter), full-wave rectified and smoothed (zero-lag 4^th^-order low-pass Butterworth filter at 7 Hz). Envelopes were normalized so that their maximum would be one. Muscle activation computations correspond to the area under the curve (AUC) for each envelope.

#### Changes on-off therapy across patients

For each participant, we computed the median value of each gait parameter across therapeutic conditions.

### Spectral and power analyses during walking experiments

#### Raw spectrograms

Spectrograms of raw LFP signals were computed through multitaper decomposition using the Chronux library (http://chronux.org/) between 5 and 125 Hz (*N* = 3 tapers, 1-s window with 50-ms overlap).

#### Average scalograms

Average scalograms were computed both for complete walking sequences (starting 2 s before the first heel off and lasting until 2 s after the last foot strike) or per gait cycle (between consecutive foot strikes of the same leg). For both cases, the scalogram of all the recorded sessions for each condition was computed using continuous wavelet decomposition (Morlet wavelets, MATLAB, USA). Each frequency was normalized with respect to the mean power computed during standing periods (as previously defined). Walking sequences were pooled together depending on whether they started with the right or left foot, interpolated (5000 points for the standing periods before and after walking and 1500 points between consecutive foot strike events during stepping), and averaged over time. For gait cycle averages, consecutive steps were pooled together after interpolation and averaged in time.

#### Computations of band power per locomotor states

For each locomotor state, we computed the power spectral density (PSD) of all the concatenated LFP corresponding to that state using the PWelsh function (one-sided, 1Hz resolution, 50% overlap). Band-power was then quantified as the area under the curve (AUC) between the frequency-limits defined for each band, once the aperiodic component 1/f from the PSD was removed. To allow direct comparisons across patients that have bands of different widths, band-power was normalised by the frequency range covered by each band.

#### Cross-correlation between band power and gait quality

Band-power and measures of gait performance were computed for each individual walking sequence, and cross-correlation coefficients were derived across all sequences (N=10 repetitions).

#### Comparisons of power per gait cycle

For each frequency band, power was computed from the bandpass filtered LFP signals (4^th^-order Butterworth filter, limits defined by the fitting algorithm). Power was normalized with respect to the mean power of all “standing” periods pooled together.

To compare band-power changes between ‘normal’ steps and ‘obstacle-avoidance’ steps, we computed the median power for each repetition of each state (N = 20 sequences) and performed non-parametric statistical analyses for each patient (Kruskal-Wallis test followed by multiple comparisons with Bonferroni correction).

### Identification of artifacted LFP channels during gait

The identification of channels corrupted by gait-related artifacts can be tricky, as both movement-related neural modulations and artifacts are locked to the rhythm of gait. Moreover, gait-related artifacts affect signals predominantly in the low-frequencies but can spread to higher frequencies, which makes them difficult to be removed through standard filtering.

To identify LFP channels that were corrupted by gait-related artifacts, we followed the same process as previously described^4^: Rather than aiming to identify artifacts in the time-domain, we reasoned that corrupted channels would exhibit important differences in the aperiodic (1/f) component of the power densities (PSD) between rest and walking: The aperiodic component captures the overall baseline power across the spectrum and should not change importantly over consecutive trials.

For each patient, we applied the fitting algorithm^1^ to the PSD of two separate recordings, one at rest (sitting) and one during walking (from the same session). We then compared their aperiodic (1/f) component: We computed the root mean square error (RMSE) in the range [10-90] Hz (region of interest) and ensured that walking did not induce an increase in 1/f power bigger than 50% compared to that sitting (difference of 1.76 dB). All channels that exceeded that value were considered as corrupted in the region of interest and discarded for further analyses or decoding purposes. Visual inspection of the spectrogram for channels labelled as “artefacted” consistently showed important low-frequency spikes that periodically corrupted the spectrogram and spread to higher frequencies. All retained channels were also verified by visual inspection of their spectrogram.

We note that this approach is highly restrictive, in that channels may still convey useful information (for instance in high frequencies that have *a priori* not been corrupted) but are nonetheless completely discarded. Out of all 63 hemispheres recoded, 2 hemispheres had to be removed during analyses during walking tasks due to movement related artetacts that strongly biased neural decoders.

### Neural decoding framework

We used a custom-built Python software (https://github.com/dbdq/neurodecode) for real-time decoding of locomotor activities from raw STN LFPs. The decoder was based on Random Forest algorithms and implemented using the python Scikit-learn library (hyperparameters: 1000 trees, 5 maximum depth, and balanced subsampling). Testing performance was computed using Monte-Carlo cross-validation over 8 repetitions (20% of data for testing, 80% for training) within the epoch windows.

Computations run on a Windows 10 at ~15 Hz (one prediction every 66 ms) as defined by computer performance. Probability traces for each class were then smoothed (alpha = 0.1) to prevent abrupt noisy peaks or artifacts. Quantification of performance is reported as the averaged F1 score per class. Feature importance vectors (contribution of each frequency band to the overall prediction) were calculated for each channel from the trained model, and normalized (sum of all contributions equal to 1).

### Genetic algorithms to optimize decoding parameters across therapeutic conditions

Neural decoders are characterized by features that define how spatial, spectral, and temporal information is to be processed from the raw neural activity. The choice of these features can markedly impact decoding performance. Defining these features manually is inherently suboptimal and biased towards a specific context (tasks, conditions), thus failing to cope with shifts in neural distributions. To prevent this bias, we developed a method based on genetic algorithms to automate the feature engineering flow (**Extended Data Fig. 7**). Our approach stochastically explored the space of potential decoders and optimizes them using a multi-objective strategy. This process allowed us to obtain a pareto-front of non-dominated decoders, i.e. those that exhibit an optimal tradeoff between the different objective functions. We employed the NSGA-II (Non-dominated Sorting Genetic Algorithm II), which is known for its robustness and efficiency in solving complex multi-objective optimization problems.

The NSGA-II operated by evolving a population of candidate decoders, each one parametrized by its set of features, or ‘genes’. Starting from a randomly-generated population of decoders, the NSGA-II algorithm applied processes inspired by biological evolution—mutation, crossover, and selection. In doing so, the population naturally evolved over successive generations. Mutation introduced variability by randomly altering genes, while crossover combined features from pairs of decoders to create offspring, potentially inheriting the strengths of each parent. The selection process was based on a ranking system where decoders are sorted into fronts based on their performance across the objective functions, primarily focusing on non-dominance and diversity. This ensured that the evolving population maintained the balance between exploring new possibilities and optimizing known good solutions, thereby efficiently navigating the complex feature space to identify optimal decoder configurations.

We first applied this automated approach to design decoders for locomotor states (sit, stand, walk) across all patients, and subsequently for L-DOPA and DBS selectors. We allowed the algorithm to evolve for at least 20 generations with a population of 50 decoders, focusing on optimizing (i) the epoch window, (ii) the range frequencies used, and (iii) the window length (the time-period within which spectral features are computed). Among all these features, window length emerged as the most significant ones for explaining decoder performance, followed by the maximum and minimum frequencies. Window lengths of [1 - 1.2] seconds provided the best robustness to decoding locomotor states, while the range [18 – 20] seconds proved optimal for the L-DOPA selector. Notably, for the locomotor decoder, this value was computed across patients, and reflects the cross-patient average dynamics that the decoder needs to exploit for good global performance. We note that these durations match the natural dynamics of the different states being decoded (gait cycles are in the range of ~1 s, whereas the absorption of L-DOPA can happen over several minutes). These values were used for the remainder of decoding computations.

### Decoding algorithms and classifiers during locomotor experiments

#### Decoder of locomotor states

Decoders were trained to discriminate between “sit”, “stand” and “walk” classes. We employed only LFP signals from contacts that were not corrupted by movement -related artifacts (see definition above). Additionally, we only considered frequencies above 10Hz to ensure that only physiological data is accounted for (range used for PSD computations = 10-100Hz, for all patients). Epoch ranges were set to 1.5 seconds (relative to the onset of each class). The onset times of each class were defined as follows: the “standing” class started at least 2s before the first step and after the last step. For the “walk” class, we divided each walking sequence in two parts and used the first. The “sit” class was defined when the patients were simply seated on a chair.

Quantification of performance (**Fig. 4,5 and Extended Data Fig. 4,5,6**) is reported as (i) median +/-sem probability traces (interpolated within consecutive footstrikes) for each class, sample-based confusion matrix across all short and long tasks, and (iii) median probability values of each class of the entire duration of each state across all short and long tasks.

#### Classifier of L-DOPA or DBS amplitude

To develop a classifier capable of distinguishing between ON and OFF L-DOPA states, or between high and low DBS amplitudes, we adapted our neural decoding framework and tuned its hyperparameters to capture therapy-induced changes in STN LFPs rather than locomotor-related modulations. Using a genetic algorithm (GA), we identified hyperparameter settings that optimally discriminated L-DOPA– and DBS-related alterations in STN LFPs (changes that occur over several minutes for L-DOPA and almost instan-taneously for DBS amplitude adjustments) while minimizing sensitivity to faster modulations linked to locomotion. With these optimized parameters, we trained separate classifiers to distinguish between (i) OFF and ON L-DOPA conditions, or between (ii) high and low DBS amplitude conditions.

### Adaptive DBS feasibility study – design

The feasibility study consisted of three experimental sessions (see timeline in **Extended Data Fig. 10a**). In Session 1 (eligibility), we assessed each participant’s cardinal motor symptoms and locomotor deficits, impedance values, as well as the quality of STN LFP signals (presence of artifacts, movement- and therapy-related modulations), both under low and high L-DOPA levels. We also assessed their response to changes in DBS amplitude. Participants meeting inclusion criteria proceeded to Sessions 2 and 3.

We applied our modular decoding framework to extract individualized neural signatures of locomotor states across varying stimulation intensities and dopaminergic states from the acquired STN LFPs. In parallel, we identified the stimulation amplitudes that optimally alleviated each participant’s most bothersome locomotor deficits under each therapeutic condition. Sessions 2 and 3 were conducted on two consecutive days, and involved the implementation of activity-dependent adaptive DBS (aDBS) protocols based on personalized neural biomarkers. We implemented these protocols on the investigational DBS platform and evaluated both the acute and sustained effects of activity-dependent DBS therapies relative to standard-of-care, continuous DBS (cDBS), both during gait assessments in- and out-of-laboratory settings. No adverse events were observed during the trial.

### Investigational DBS platform

The investigational Medtronic aDBS system used in this study was composed of a CE-marked Percept PC neurostimulator (Model B35200) and Sensight DBS leads (Models B33015), connecting to a Clinician Mobile Platform (Model CT900) endowed with an investigational DBS clinician programmer application (A610, v3.0).

This platform allowed continuous, real-time monitoring of STN LFP power for a single, user-defined frequency band (5Hz band around a selected frequency value) selected in the range 7.8Hz-70Hz. Band power was calculated by applying a Fast Fourier Transform (FFT) to a predefined time-domain window, with spectral estimates averaged across overlapping windows (80% overlap)^5,6^. These band-power modulations were used as control signal to trigger corrections in DBS parameters (up- or down-regulation of stimulation amplitude) based on one or two configurable thresholds. Three adaptive DBS modalities were available^5^:

i. Dual-Threshold mode: Stimulation amplitude is increased when LFP power exceeds the upper threshold, decreased when it drops below the lower threshold, and remains unchanged when power is within range.
ii. Single-Threshold mode: Stimulation amplitude is increased when LFP power exceeds the threshold and returned to the lower set amplitude when it goes below the threshold. This mode was not used in the present study, as it requires calibration at 0 mA stimulation, posing challenges during motor tasks for most of the participants.
iii. Single-Threshold Inverse mode: Stimulation amplitude is decreased when LFP power exceeds the threshold, and returned to the higher amplitude when it goes below the threshold.

Overall, this investigational platform allowed access to features not available in the CE-marked clinical version. These included: (i) use of LFP gamma band power (30–70 Hz) for control of stimulation, in addition to alpha (7–10 Hz) and beta (10–30 Hz) bands; and (ii) selection of the single-threshold inverse aDBS mode.

### Identification of patients’ most bothersome gait deficits

Each participant’s most disabling gait impairments were identified during Session 1 based on clinical reports, subjective feedback, and in-lab clinical assessments of gait performance (Timed Up and Go test, 360° turn test, MDS-UPDRS Part III, and tailored freezing-of-gait circuit), and quantitative movement analyses performed under both low and high dopaminergic states by a movement disorders specialist (**Extended Data Table 3**). Similar to experiments to characterize the physiological principles of activity-dependent encoding (part 1), we assessed gait deficits using synchronized bilateral EMG recordings and whole-body kinematics during a comprehensive set of locomotor tasks (**Extended Data Fig. 9a**). Gait quality was further quantified using a camera-based motion tracking system (OpenCV^7^ and RTMPose^8^) that extracted metrics of posture, balance, and interlimb coordination, serving as a portable alternative to traditional optoelectronic motion capture.

### Identification of optimal DBS amplitudes to address gait deficits

We first verified the range of tolerated stimulation amplitudes for each hemisphere. Tolerability to dynamic changes in amplitude within that range was further assessed using the cycling stimulation mode, in which the implantable neurostimulator was configured to toggle stimulation ON and OFF at two predefined rates (1s and 50 ms). Participants then performed a series of motor tasks customised to expose their gait deficits, e.g. walking back-and-forth and ambulation in a FOG circuit, under four stimulation conditions: (i) the standard-of-care amplitude, (ii) the maximal amplitude and minimal amplitudes within the therapeutic window, and (iii) 0 mA (when tolerated). Based on motor performance across these conditions, a movement disorders specialist identified the stimulation amplitude that most effectively alleviated each participant’s predominant gait impairment.

### Identification of personalized neural signatures

LFP, EMG, and kinematic data were analyzed offline to: (i) assess neural signal quality, including the presence of artifacts such as ECG contamination or movement-induced noise that could interfere with aDBS implementation, and verify acceptable electrode impedance; and (ii) evaluate the consistency of LFP modulations associated with locomotor activity across two stimulation amplitudes optimized for different motor symptoms (i.e., cardinal and gait-related).

To identify the personalised neural signatures, we applied our decoding framework on LFP recordings acquired during motor tasks performed at the two stimulation amplitudes. The training dataset for the decoding algorithm consisted of 30-second periods of rest (sitting) and walking back and forth. For participants unable to perform sustained walking due to fatigue, continuous leg movements were used as a simplified task. Periods of movement and rest were segmented using EMG and kinematic signals. The tasks were repeated in both high and low dopaminergic states. For participant P1, the low dopaminergic condition was not analyzed due to motor impairment preventing completion of the necessary clinical evaluations.

Spectral features were extracted in the 7–70 Hz range, as permitted by the constraints of the DBS platform for real-time control. The lower bound was increased to 15 Hz in cases where low-frequency components raised concerns about walking-related artifacts. Decoding analyses were conducted separately for each dopaminergic state. For each condition, features were extracted unilaterally, and decoders were trained independently for the left and right hemispheres. This approach allowed to identify the most informative spetral features per hemisphere to discriminate between locomotor and resting states across different dopaminergic states and stimulation amplitudes.

We finally confirmed visually that movement-related modulations in the identified frequency bands corresponded to a synchronization or a desynchronization relative to the resting state.

### Configuration of activity-dependent adaptive DBS therapy

We then leveraged these hemisphere- and therapy-specific neural features to configure activity-dependent DBS for each participant. **Extended Data Fig. 9** shows the workflow followed to configure the adaptive therapy.

The optimal frequency band for each hemisphere was selected from among the spectral features suggested by the decoders. When multiple candidate spectral features were found to be informative, they were ranked according to their contribution, evaluated during online assessments, and selected based on real-time reliability. If similar informative spectral features emerged across dopaminergic conditions, the same frequency band was selected for both conditions to enhance the stability of real-time adaptations. In cases where one hemisphere provided more robust decoding performance than the contralateral hemisphere, a unilateral control signal was tested to control the adaptions of bilateral DBS amplitudes.

In two participants, we found cross-session variations in the stability of the most informative features due to day-to-day variations. The hemisphere- and frequency-band used for DBS adaptations were tuned based on the previous ranking to maximise control robustness across these sessions.

The choice of aDBS mode was dictated by the need to either up- or down-regulate stimulation amplitude during locomotion, relative to the standard-of-care setting optimized for cardinal motor symptoms, in order to deliver the amplitude that best alleviated gait impairments. **Extended Data Fig. 10** summarizes the selected control frequency bands and whether unilateral or bilateral hemispheres were monitored for DBS control.

Threshold tuning was performed using the dedicated tablet interface. To enable robust discrimination of locomotor activities using a single threshold (and minimize instabilities during adaptive changes in DBS amplitude), LFP recordings for each activity were acquired at distinct stimulation amplitudes. Specifically, participants performed 30 seconds continuous walking at their standard-of-care DBS amplitude, followed by a 30-second rest period at the amplitude that best alleviated their gait deficits. In participants unable to perform repeated walking trials, walking was substituted with continuous seated leg movements. This pairing of task and stimulation amplitude was designed to enhance contrast in LFP power between states, thereby improving the reliability of threshold calibration. During these tasks, the DBS platform computed LFP band power in real time and automatically computed threshold values. These values were then manually refined based on analyses from Session 1 and visual inspection of the consistency of LFP modulations relative to the threshold online testing.

Stimulation contact configuration, frequency, and pulse width remained constant across all experimental sessions and conditions. **Extended Data Table 4** provides a detailed overview of the final aDBS parameters used to target each participant’s gait impairment.

### In-lab assessment of acute effects

To evaluate the acute effects of activity-dependent DBS, we selected clinical tests and motor tasks tailored to each participant’s most disabling gait impairment. Tasks were performed under both aDBS and cDBS, with conditions randomly administered across repetitions:

- Participant P1: Performance was assessed using the 5-meter Timed Up and Go (TUG) test, repeated three times per condition, to capture their reduced leg agility impairing flexion of the right flexion and gait fluidity (**Extended Data Fig. 11a)**. The test was repeated three times per condition.
- Participant P2: The main deficit was freezing of gait, which occurred predominantly during turning, passing through doorways, and obstacle avoidance. We assessed this using the 360-degree turn test (8 repetitions) and a customized freezing-of-gait circuit comprising three freezing-prone locations (**Extended Data Fig. 11c**). Each circuit was repeated twice per condition.
- Participant P3 reported both FOG and foot dyskinesia. Performance was evaluated using repeated trials of an 8-shaped FOG circuit^9^ and three back-and-forth walking trials per condition.

LFP and biomechanical signals were recorded during all tasks using our multimodal monitoring platform. FOG episodes were identified from video recordings by a movement disorders specialist. FOG episode durations were quantified using EMG analysis, and turning sequences were identified by gyroscope signals. Gait quality was assessed both by a movement disorders neurologist and through quantitative EMG and kinematic parameters.

### Out-of-lab assessments

To evaluate the sustained efficacy and robustness of activity-dependent DBS in daily life conditions, participants performed tasks in out-of-laboratory settings during Session 3 under both adaptive DBS and standard-of-care continuous DBS. Stimulation conditions were randomly assigned and remained blinded to participants until the session was completed.

The out-of-laboratory assessments in participant P2 included walking along a path within the hotel where the participant stayed overnight between sessions 2 and 3. This environment was chosen to replicate a naturalistic setting (**Extended Data Fig. 11d**). Participant P3 was instructed to walk outdoors and retrieve a snack from the hospital Iteria. In both cases, a member of the research team accompanied the participant at all times to ensure safety and allow for real-time monitoring.

LFP and kinematic data were acquired using sensorized shoes with embedded machine learning algorithms that detect walking events and compute gait parameters in real time (NUSHU Research, Magnes, Swizterland). These were aligned with STN LFP recordings. For participant P2, a portable EMG system (Delsys, USA, operating at 1.3kHz) was additionally used to record activity from two leg muscles (left vastus medialis VM, and tibialis anterior TA) to quantify FOG duration and two IMU sensors (sampling frequency: 370Hz) were placed on each foot to identify turning frequency. The aDBS system functioned reliably throughout these real-world tasks. Analyses of kinematic and neural data were conducted offline following the experiments to compare gait quality between activity-dependent DBS and standard-of-care continuous DBS.

Participant P1 was unable to complete the out-of-laboratory evaluation due to excessive fatigue following the optimization of aDBS therapy and in-laboratory assessments.

To assess participants’ subjective experience with activity-dependent DBS in daily life, each individual completed a custom-designed questionnaire. They rated the perceived impact of adaptive DBS on various Parkinsonian symptoms (gait, FOG, tremor, balance, rigidity, dyskinesia) using a scale from –10 (much worse) to +10 (much better), relative to their standard-of-care stimulation.

### Data Analysis and Statistical Methods

EMG and gyroscope data recorded under activity-dependent DBS and standard-of-care continuous DBS were analyzed using Statistical Parametric Mapping (SPM1D, https://spm1d.org). For each muscle, EMG envelopes were extracted for individual gait cycles and time-normalized. An unpaired two-tailed t-test was then used to assess statistical differences in EMG activity across gait cycles between stimulation conditions. The same analysis was applied to foot angular velocity derived from gyroscope signals.

In-laboratory gait quality was monitored during unconstrained locomotion using camera-based motion tracking algorithms (OpenCV^7^ and RTMPose^8^) developed in Python. To extract leg, arm, and trunk parameters, kinematic signals was discretised per gait-cycle, parametrised into 77 discrete parameters (e.g., joint and elevation angles and velocities for each leg and arm joint, and trunk, as well as the relative timing of stepping phases), zscored, and projected into a lower dimensional space using principal component analysis for visualisation, both under adaptive DBS and continuous DBS. Five healthy controls were included in PCA analyses for direct comparisons of changes in gait quality.

During out-of-laboratory assessments, gait parameters were extracted using the NUSHU online platform^10^ (Magnes, Switzerland) and compared between adaptive DBS and continuous DBS using unpaired two-tailed t-tests. These analyses were conducted in MATLAB (MathWorks, Natick, MA, USA). A significance level of alpha equal to 0.05 was applied for all statistical tests. Bonferroni correction was applied for multiple comparisons.

### Statistics

Experimental data and simulations were processed offline using MATLAB R2024a (MathWorks). All data are reported as mean or median values, ±sd or sem, as indicated. Normality of data was tested using a Kolmogorov-Smirnov test with 95% confidence interval (CI). Comparisons between conditions were computed using one-tailed t test (parametric) or Wilcoxon sign rank test (non-parametric) with 95% CI. Multi-group comparisons were performed using one-tailed ANOVA (parametric), or Kruskal-Wallis (non-parametric) tests with 95% CI, followed by Bonferroni correction.

## DATA & CODE AVAILABILITY

All data associated with this study are present in the paper or the Extended Data Materials. Anonymized data that support the findings of this study are available in this Zenodo DOI (link to be included). Custom code used in this study is available from the same Zenodo DOI (link to be included).

## ACKNOWLEDGEMENTS

This work was funded by the Swiss National Science Foundation (Grant number TMSGI3_218471), the Foundation for the Advancement of Neurology (project NeuroGAIT), the Parkinson Move Foundation, and the Defitech Foundation. The study was supported by Medtronic Inc. through the provision of the investigational Percept Tablet. Medtronic had no role in the design of the study, data collection, analysis or interpretation, or in the writing of the manuscript.

We thank Loïc Comeliau and Maxime Baud for their contributions to the development of the multimodal recording platform and for their support during the experimental sessions. We also thank Alessandro Schaer and George Chatzipirpiridis for their support with data export and analysis of the NUSHU sensorized shoes.

## AUTHOR CONTRIBUTION

S.S, V.d.S., R.W, C.V., G.C., J.B., and E.M.M. conceived the study. S.S., V.d.S., R.W., P.S.L, C.V., Y.T. and E.M.M. established technological platforms and acquired data to characterize physiological principles of locomotor encoding within STN dynamics. V.d.S., C.V., L.B-F. and E.M.M wrote the feasibility clinical protocol to deliver activity-dependent adaptive deep brain stimulation. V.d.S., R.W., C.V., C.D. and E.M.M established the platform and acquired data on activity-dependent adaptive deep brain stimulation. S.S, V.d.S., R.W, P.S.L., I.E., A.S.L and E.M.M performed electrophysiological and biomechanical data analyses. S.S., I.S., and K.L built decoding algorithms and analyzed their performance. N.B. performed anatomical reconstructions of DBS lead placement. M.B. and F.M. designed three-dimensional illustrations and helped with figure editing. C.V., C.D., V.F., E.A., B.W., C.H., M.C.J., J.F.B., and J.B. recruited patients and performed clinical evaluations. J.B. performed surgeries. S.S, V.d.S., R.W. P.S.L, C.V., I.S., G.C., J.B. and E.M.M. prepared the figures. R.W. and E.M.M. prepared the supplementary videos. S.S., V.d.S., G.C., J.B., and E.M.M. wrote the manuscript. All authors contributed to its editing. A.P., H.L., A.C.C, G.C., J.B. and E.M.M. acquired funding. E.M.M. supervised all aspects of the work.

## ADDITIONAL INFORMATION

**Extended Data Figure 1** | Anatomical reconstructions of deep brain stimulation lead placement across participants.

**Extended Data Figure 2** | Electrophysiological characteristics of STN local field potentials across participants.

**Extended Data Figure 3** | Therapy-specific modulations in activity-dependent STN dynamics

**Extended Data Figure 4** | Participant- and therapy-specific neural decoders are required to maximize prediction performance within therapeutic conditions

**Extended Data Figure 5** | Decoding performance of locomotor states across participants.

**Extended Data Figure 6** | Modularity preserves decoding accuracy across therapeutic conditions.

**Extended Data Figure 7** | Genetic algorithm to optimize the parameters and hyper-parameters of neural decoders and selection logic.

**Extended Data Figure 8** | Out of laboratory performance of modular decoder

**Extended Data Figure 9** | Methodology to setup activity-dependent adaptive DBS

**Extended Data Figure 10** | Patient-specific tuning of activity-dependent DBS based on individualized physiological decoding

**Extended Data Figure 11** | Patient-specific Improvements in locomotor deficits mediated by activity-dependent DBS

**Extended Data Table 1** | Demographic and clinical characteristics of all participants included in the characterization of physiological principles underlying therapy-specific encoding of locomotor activities within STN dynamics, and the development of neural decoders.

**Extended Data Table 2** | Locomotor tasks and therapeutic conditions recorded in all participants included in the characterization of physiological principles underlying therapy-specific encoding of locomotor activities within STN dynamics.

**Extended Data Table 3** | Demographic and clinical characteristics of the seven participants recruited for the proof-of-concept feasibility study assessing the safety and preliminary efficacy of activity-dependent adaptive DBS.

**Extended Data Table 4** | STN DBS parameters used for activity-dependent adaptive DBS targeting locomotor deficits, compared to standard-of-care continuous DBS.

## SUPPLEMENTARY MATERIALS

**Supplementary Video 1** | Physiological principles of activity-dependent STN dynamics across therapeutic conditions

**Supplementary Video 2** | Modular decoding framework to predict locomotor activities across L-DOPA fluctuations

**Supplementary Video 3** | Activity-dependent DBS therapies alleviate locomotor deficits and cardinal motor symptoms

## Extended Data

**Extended Data Figure 1.**
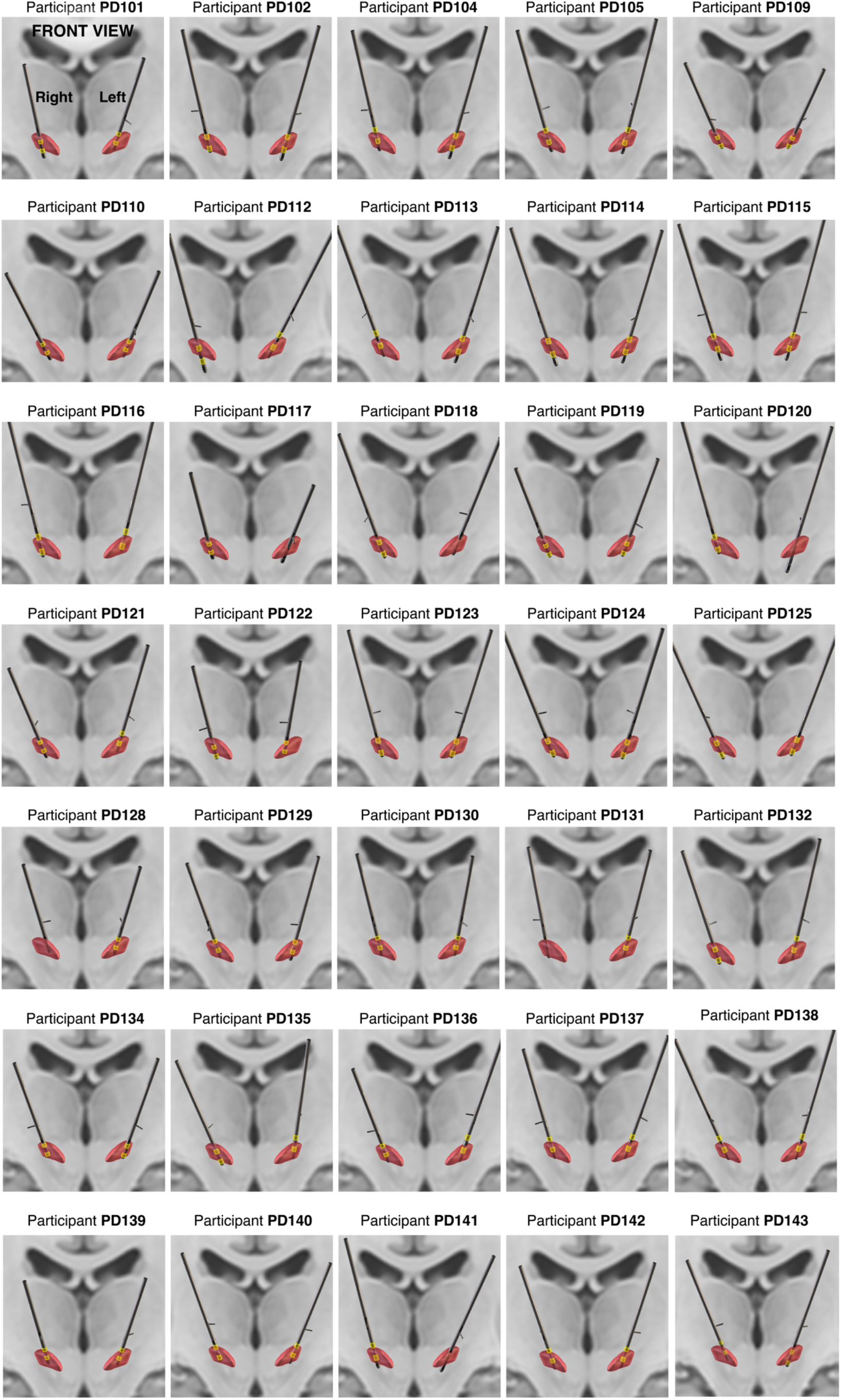
Anatomical reconstructions of deep brain stimulation lead placement across participants. Bipolar contacts used for STN LFP recordings are highlighted in yellow for each hemisphere.

**Extended Data Figure 2.**
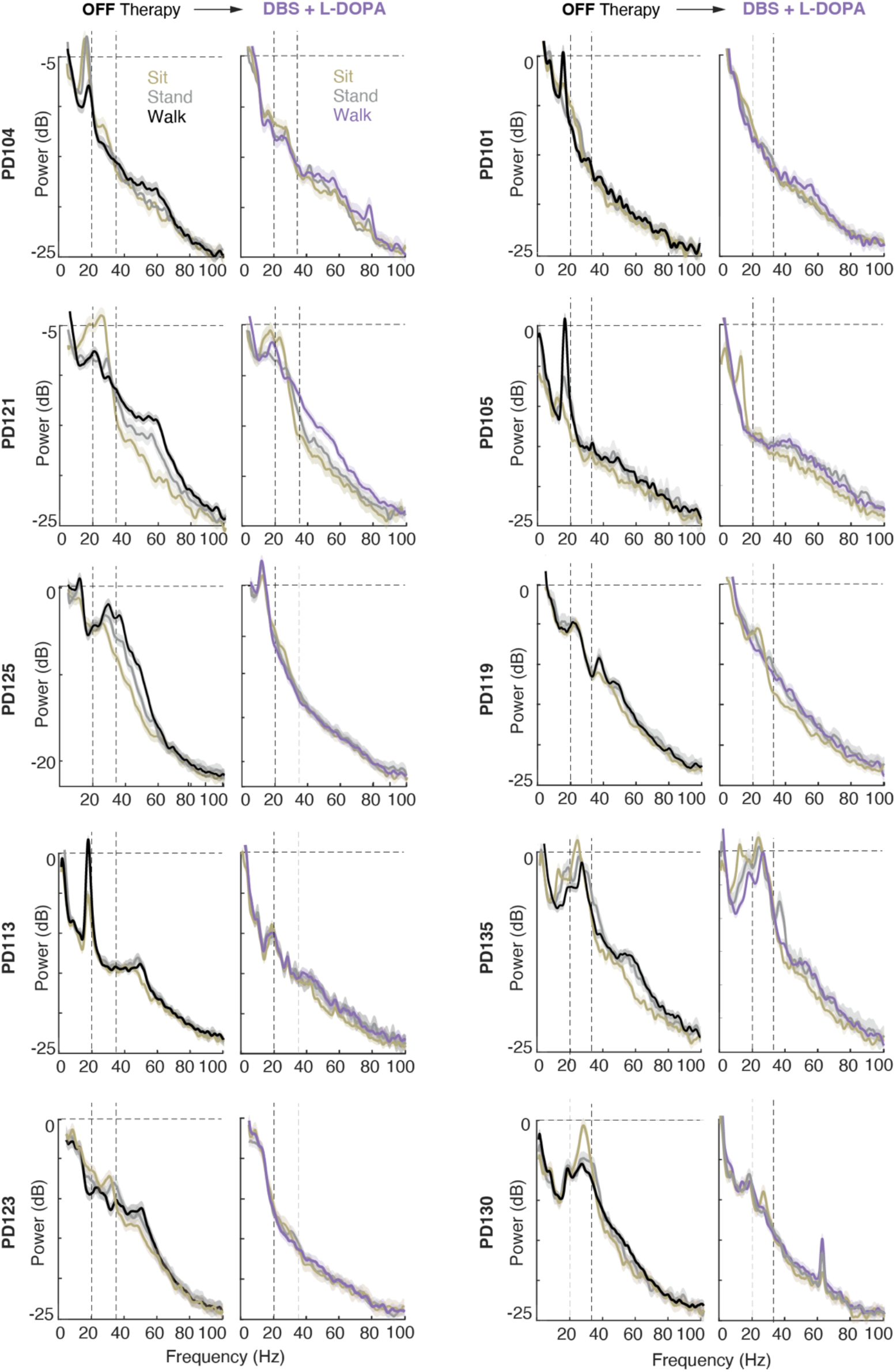
Electrophysiological characteristics of activity-dependent modulations within STN dynamics across 10 illustrative participants, shown both in their most parkinsonian state (OFF therapy) and under their standard-of-care combination of STN DBS and L-DOPA. For each participant, power spectral densities are shown for each activity. Dashed lines highlight the boundaries of the high-beta canonical band (20-35 Hz). Several of these participants are featured in later figures to illustrate activity- and therapy-specific modulations. Comparison under standardized conditions is intended to facilitate interpretation of the individualized adaptations presented throughout the manuscript.

**Extended Data Figure 3.**
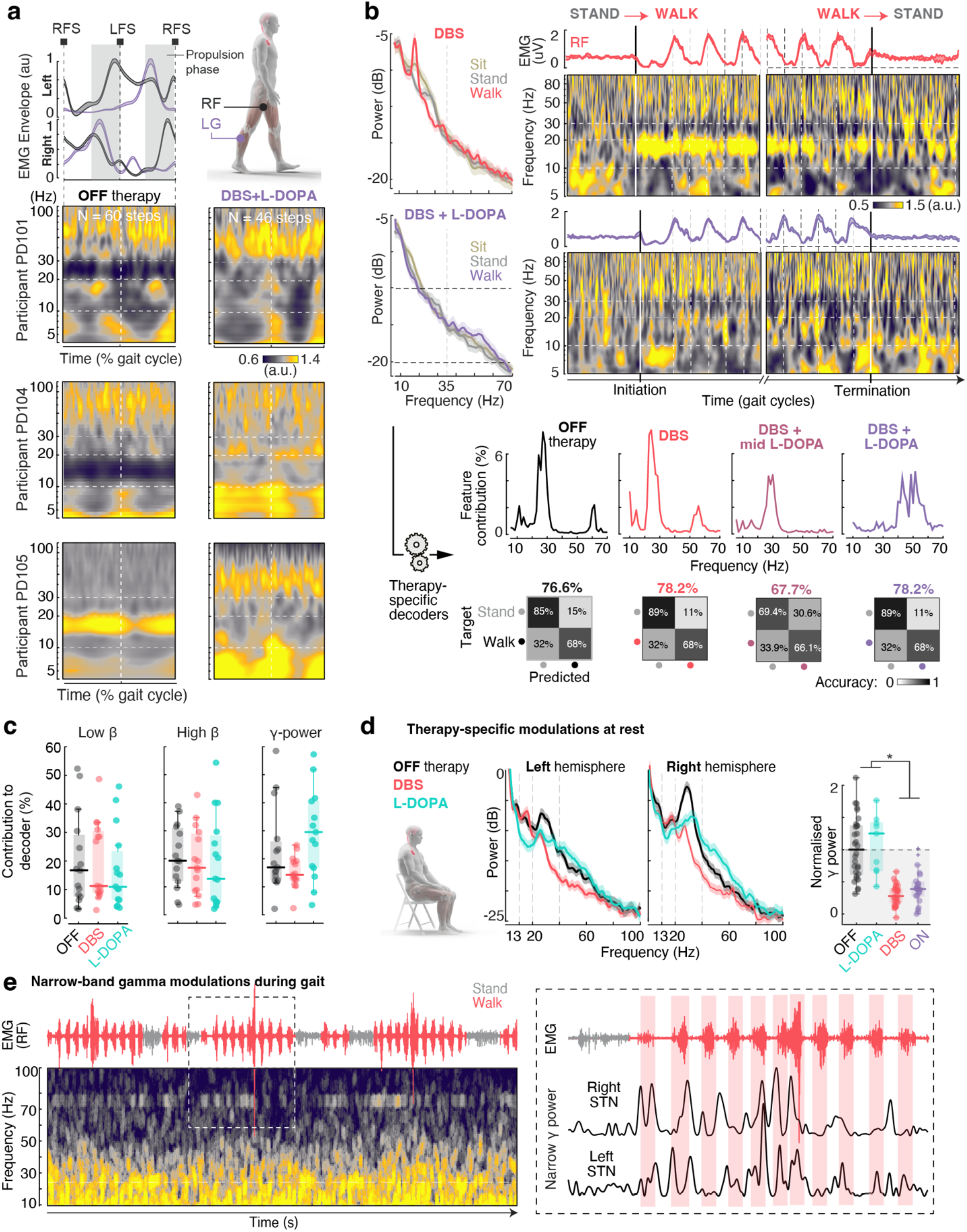
Therapy-specific modulations in activity-dependent STN dynamics. **(a)** Representative examples of averaged (mean ± SEM) EMG envelopes for two leg muscles (Rectus Femoris [RF] and lateral Gastrocnemius [LG]), interpolated between contralateral foot strikes (Right FS and Left FS), and temporally aligned to averaged time-frequency scalograms of STN LFPs from three different participants (PD101, PD104, and PD105, top to bottom). Spectral power is normalized to the standing state and shown under two conditions: OFF therapy and the standard-of-care combination of L-STN DBS and L-DOPA. Patients exhibit gait-phase-specific power modulations. In the OFF-therapy condition, these modulations include bursts of low-beta synchrony during the propulsion phase (aligned with LG activation), which fade under standard-of-care therapy and are replaced by gamma-band modulations. However, marked inter-individual differences in gait encoding are evident. **(b)** *(Top*) Power spectral density (PSD) of activity-dependent STN modulations and corresponding average time-frequency scalogram across repeated sequences of stand–walk–stand (N = 10), aligned to the initiation and termination of walking (participant PD101). Two conditions are shown: STN DBS only and the standard-of-care combination of L-DOPA and STN DBS. (*Bottom*) Contribution of spectral features to neural decoders distinguishing standing from walking, illustrating the gradual shift in gait-related encoding across conditions. Confusion matrices summarize classification performance under each condition. **(c)**Therapy-specific contributions of each frequency band (low beta, high beta, or gamma) to neural decoders predicting locomotor movement from rest across N = 15 participants. Decoders trained in the OFF-therapy condition rely more heavily on low-beta activity compared to those trained under STN DBS alone or under L-DOPA alone. In contrast, decoders trained under L-DOPA rely more strongly on gamma activity, reflecting the shift in gait encoding observed during the characterization of activity-dependent modulations within STN dynamics. **(d)** Distinct band-specific modulations mediated by each therapy are also observed during rest. Boxplots quantify changes specifically in the gamma band. **(e)** Illustrative example of raw EMG trace for one leg muscle (Rectus Femoris RF) aligned to raw spectrogram of STN LFPs, showing modulations in narrow gamma band power (emerging at half the stimulation frequency) during a locomotor task involving interleaved periods of standing, walking, and obstacle avoidance under the standard-of-care therapy. The inset zooms into one walking sequence and displays bilateral traces of narrow gamma band power, which exhibit modulations time-locked to individual steps.

**Extended Data Figure 4.**
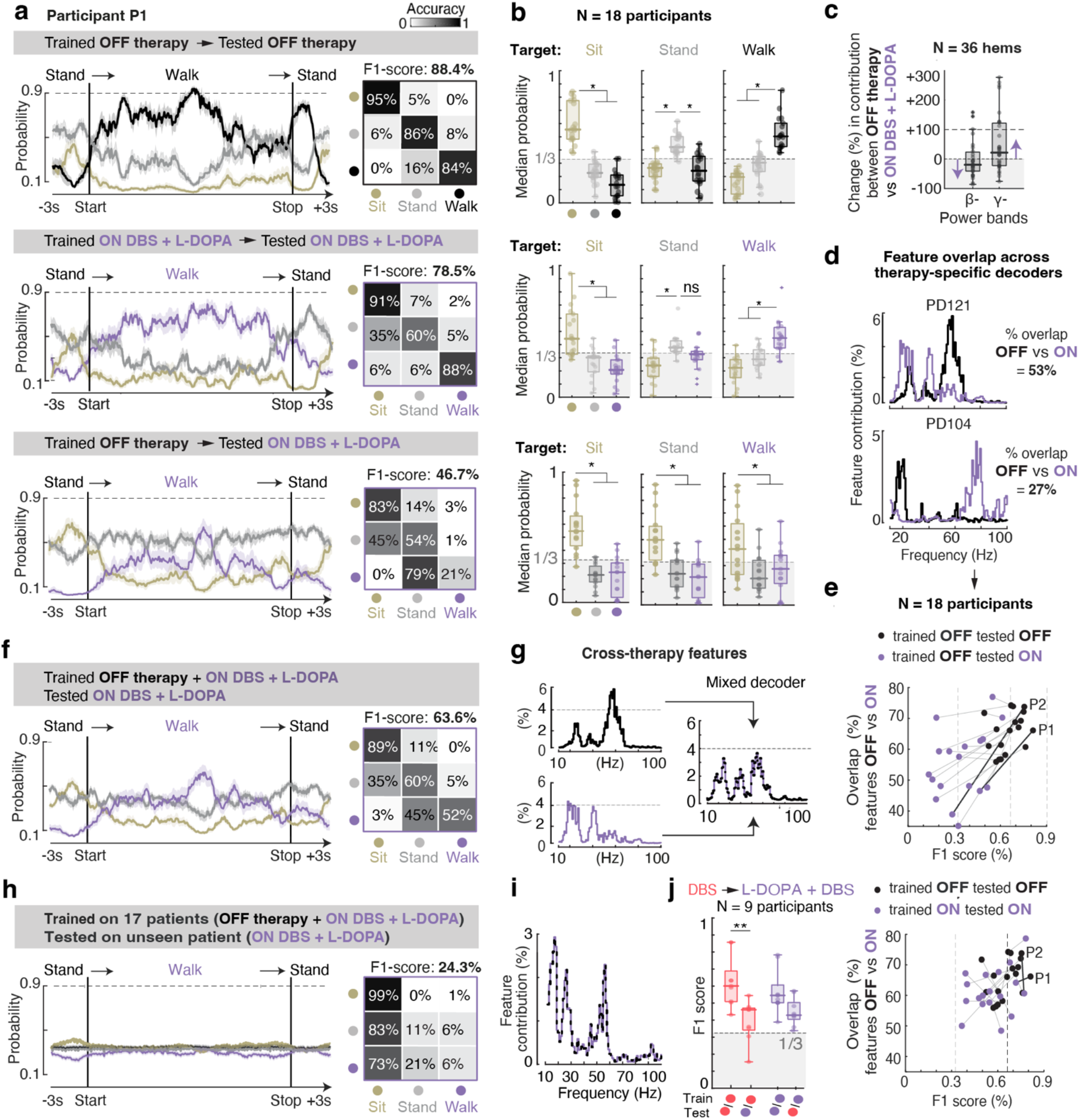
Therapy-specific neural decoders are required for optimal performance within therapeutic conditions. **(a)** Representative examples of average probability traces for three decoded activities (stand → walk → stand) during a complete walking sequence (participant PD121). Decoders were trained and tested either within the same or across different therapeutic conditions. (*Right*) Correspondoing confusion matrices and F1-scores for each decoder. **(b)** Boxplots of mean probability values for each class across all 18 participants, shown for the same three conditions as in (a). Therapy-specific decoders accurately classify each activity, whereas cross-therapy decoding exhibits systematic bias toward incorrect classes. **(c)** Comparison of beta (13–35 Hz) and gamma (50–90 Hz) band contributions to therapy-specific decoders. Across all 36 hemispheres, decoders trained under the standard-of-care combination of STN-DBS + L-DOPA rely more heavily on gamma power and less on beta power than decoders trained in the OFF-therapy condition. This pattern is consistent with the modulation observed in these bands under standard-of-care therapy. However, individual feature contributions vary substantially across participants, reflecting the unique spectral encoding of locomotor states in each case. **(d)** The transferability of neural decoders across therapeutic conditions depends on the degree of feature overlap between conditions Two illustrative examples showing the overlap in spectral features between decoders trained in the OFF-therapy state and under the standard-of-care combination of STN-DBS + L-DOPA. **(e)** Correlation between the degree of feature overlap and decoding performance within and across therapeutic conditions. **(f)** Training decoders on combined data from both therapeutic conditions yields variable performance, with a persistent bias toward incorrect classifications. **(g)** Contribution of spectral features to therapy-specific versus cross-therapy decoders in participant PD121. Decoders trained across both therapeutic conditions distribute weights more broadly across frequency bands, reducing spectral specificity and overall classification performance. **(h)** Illustrative example of average probability traces and confusion matrix for a cross-therapy decoder trained on data from 17 participants and tested on an unseen participant. Predictive accuracy displays chance-level performance. **(i)** Spectral feature contributions across participants reveal three dominant frequency bands (low-beta, high-beta, and gamma), consistent the spectral components commonly associated with movement encoding in patients with Parkinson’s disease. However, this generalized approach fails to capture participant-specific modulations critical for accurate decoding. **(j)** Cross-therapy performance of decoders trained and tested under STN DBS alone versus the standard-of-care combination of STN DBS and L-DOPA.

**Extended Data Figure 5.**
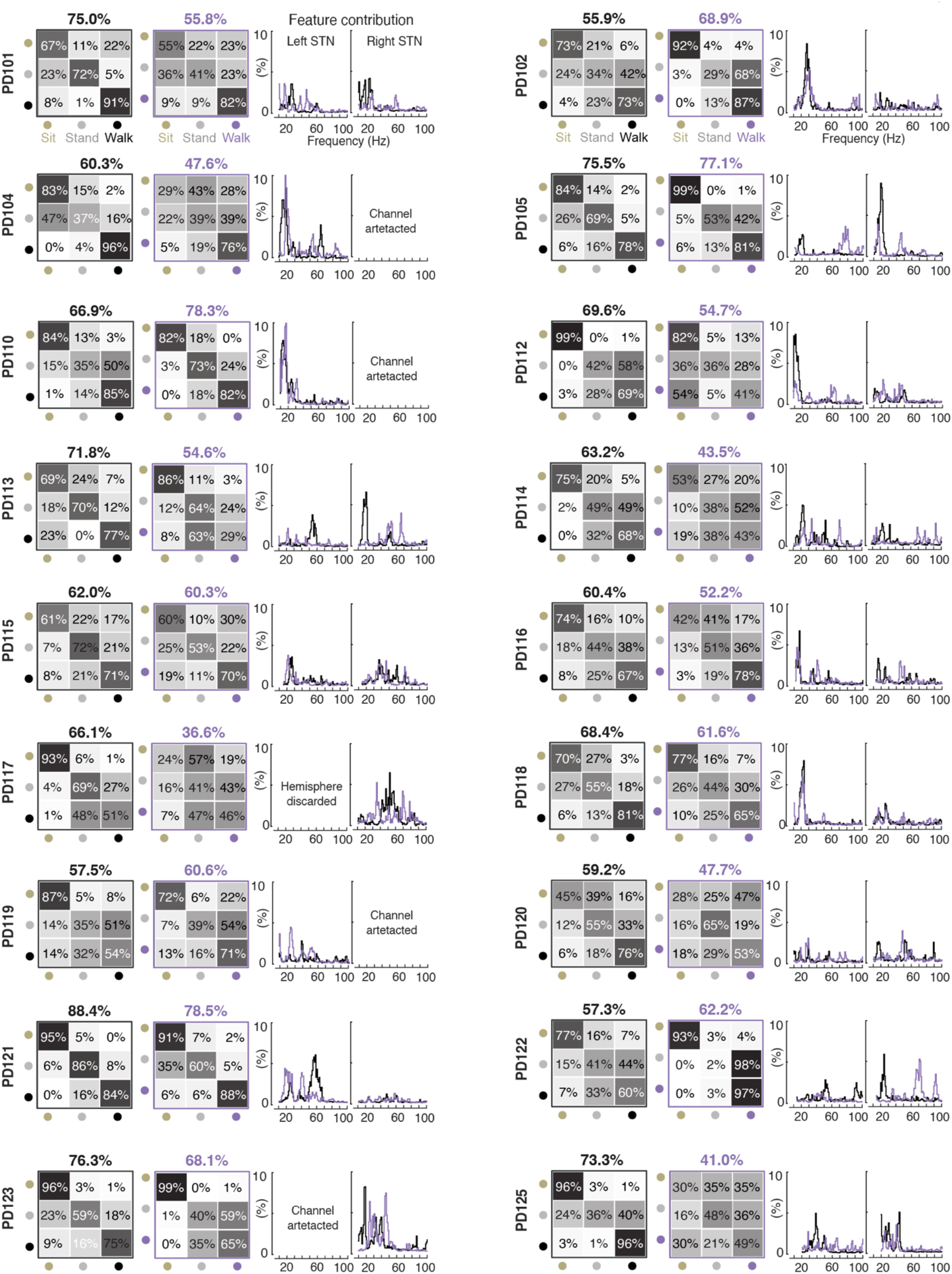
Participant-specific decoding performance of locomotor states under each therapeutic condition. Confusion matrices and spectral feature contributions for each decoder, shown by participant and therapeutic condition (OFF-therapy: black; Standard-of-care combination of STN-DBS + L-DOPA: purple). Channels excluded due to artifacts or suboptimal electrode placement are indicated accordingly.

**Extended Data Figure 6.**
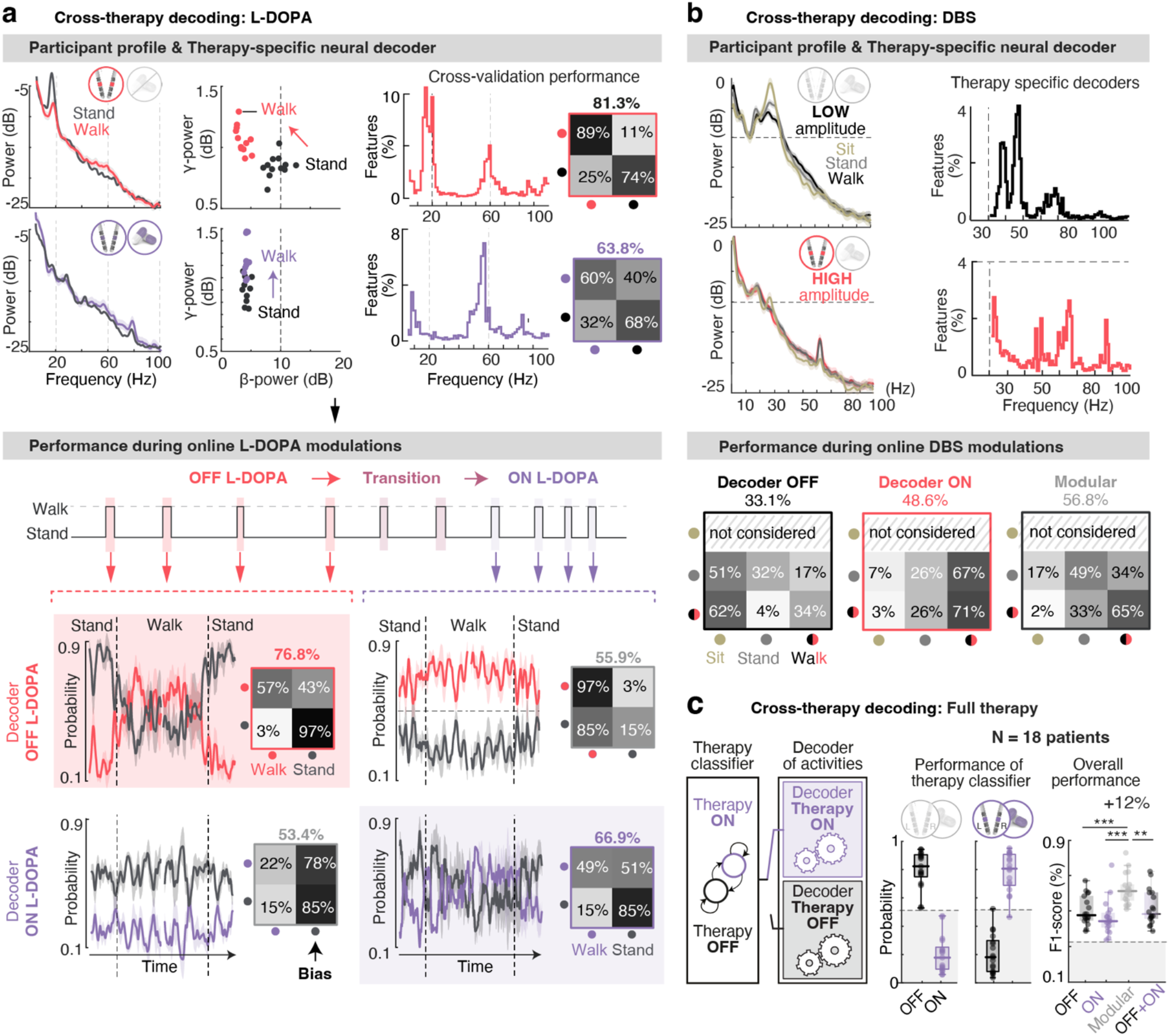
A modular decoding framework improves accuracy and generalization across therapeutic conditions. **(a) (***Top*) Changes in activity-dependent STN dynamics mediated by L-DOPA in participant PD104 (displayed in **Fig. 5a-e**). The power spectral density and epoch distributions in feature space are presented for two activities (stand and walk), alongside the predictive performance of therapy-specific decoders and the corresponding neural features identified for each therapeutic condition. (*Bottom*) Decoder performance during online L-DOPA fluctuations, illustrating the progressive change in classification accuracy as L-DOPA is absorbed. **(b) (***Top*) Changes in activity-dependent STN dynamics mediated by differences in DBS amplitude (0 mA vs. standard-of-care amplitude) in participant PD130 (shown in **Fig. 5f-i**), alongside the neural features identified for each therapeutic condition. (*Bottom*) Performance of each therapy-specific decoder during a recording in which DBS amplitude was manually toggled between the two settings, illustrating the progressive change in classification accuracy as L-DOPA is absorbed. **(c)** The modular decoding framework maintains prediction accuracy when generalizing across the most distinct therapeutic conditions (OFF therapy and standard-of-care STN-DBS plus L-DOPA).

**Extended Data Figure 7.**
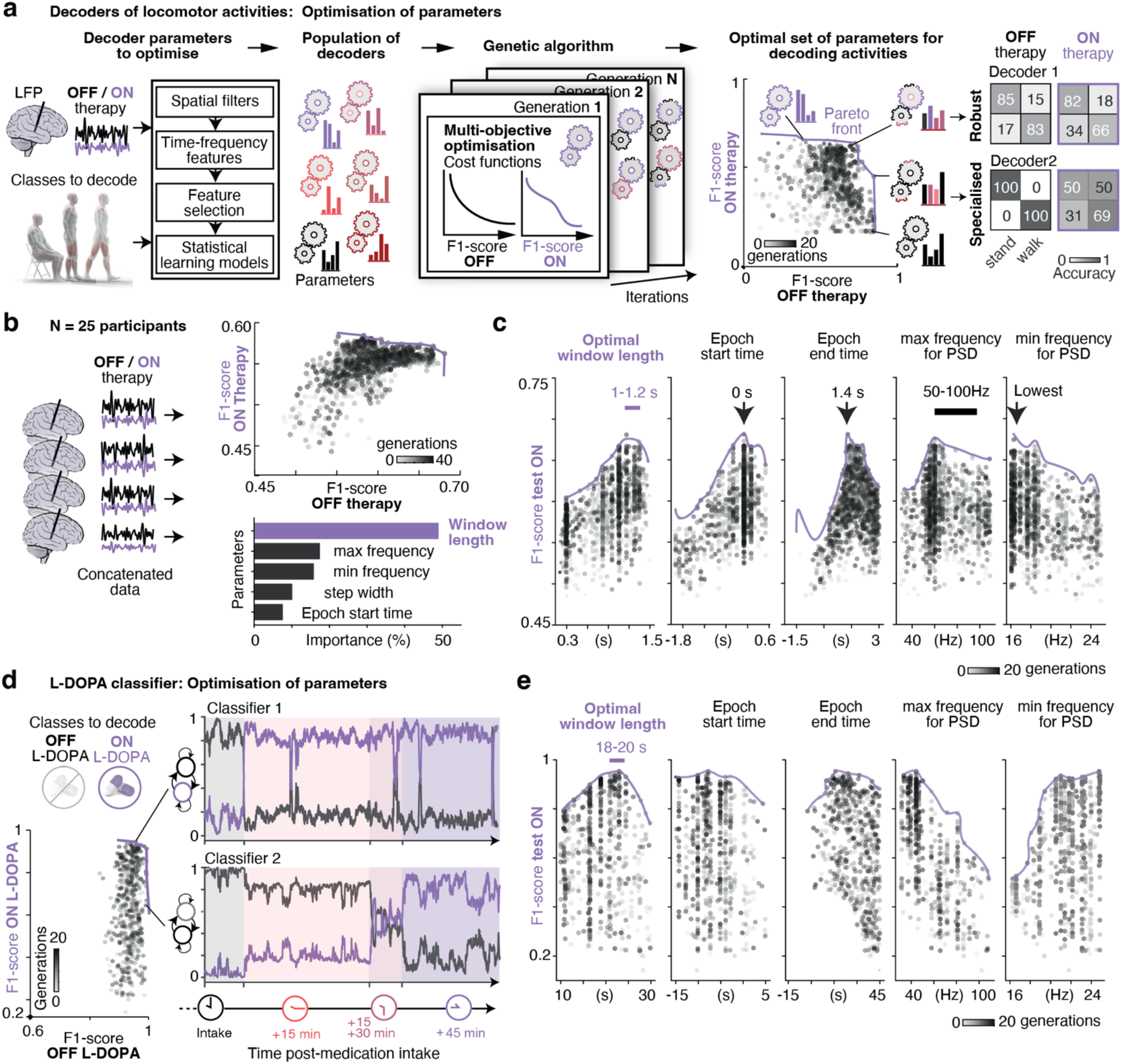
Genetic algorithm optimizes hyper-parameters of neural decoders and therapy classifiers. **(a)** Schematic of the multi-objective optimization pipeline using genetic algorithms to automate feature selection. An initial population of 50 decoders with different parameters is iteratively evolved through mutation, crossover, and selection over 20 generations to maximize model performance based on a predefined fitness criteria. The final output forms a Pareto front of non-dominated solutions balancing competing objectives, namely accuracy and robustness across therapeutic conditions. Confusion matrices illustrate the performance of two representative decoders along this Pareto front: Decoder 1 prioritized robustness across therapeutic conditions, whereas Decoder 2 was optimized for accuracy within one condition. **(b)** Application of the pipeline to identify and optimize key parameters for decoding locomotor activities (sit, stand, walk) across participants. Parameters optimized include: (i) the epoch window, (ii) the frequency range, and (iii) the window length used for spectral feature extraction. Feature importance analysis revealed window length as the most influential parameter, followed by the upper and lower frequency bounds. **(c)** Scatter plot of parameter values across generations for all assessed features. The optimal value maximizing decoding performance is indicated for each parameter. Window length values consistently converged in the range of 1.0–1.2 seconds. This duration matches the natural dynamics of different activities being decoded (gait cycles are in the range of ~1 s, and standing phases lasted a few seconds during our experiments) **(d)** Application of the same pipeline to identify and optimise L-DOPA therapy classifiers. Two representative classifiers illustrate optimized parameters for accuracy (Classifier 1) and robustness (Classifier 2). **(e)** Optimal window lengths for L-DOPA classifiers ranged between 18–20 seconds, consistent with the slower pharmacokinetics of L-DOPA absorption. The differential tuning of parameters for activity decoders versus therapy classifiers leverages the distinct temporal dynamics of locomotion and therapy-induced effects to disentangle their respective contributions from the same neural signal.

**Extended Data Figure 8.**
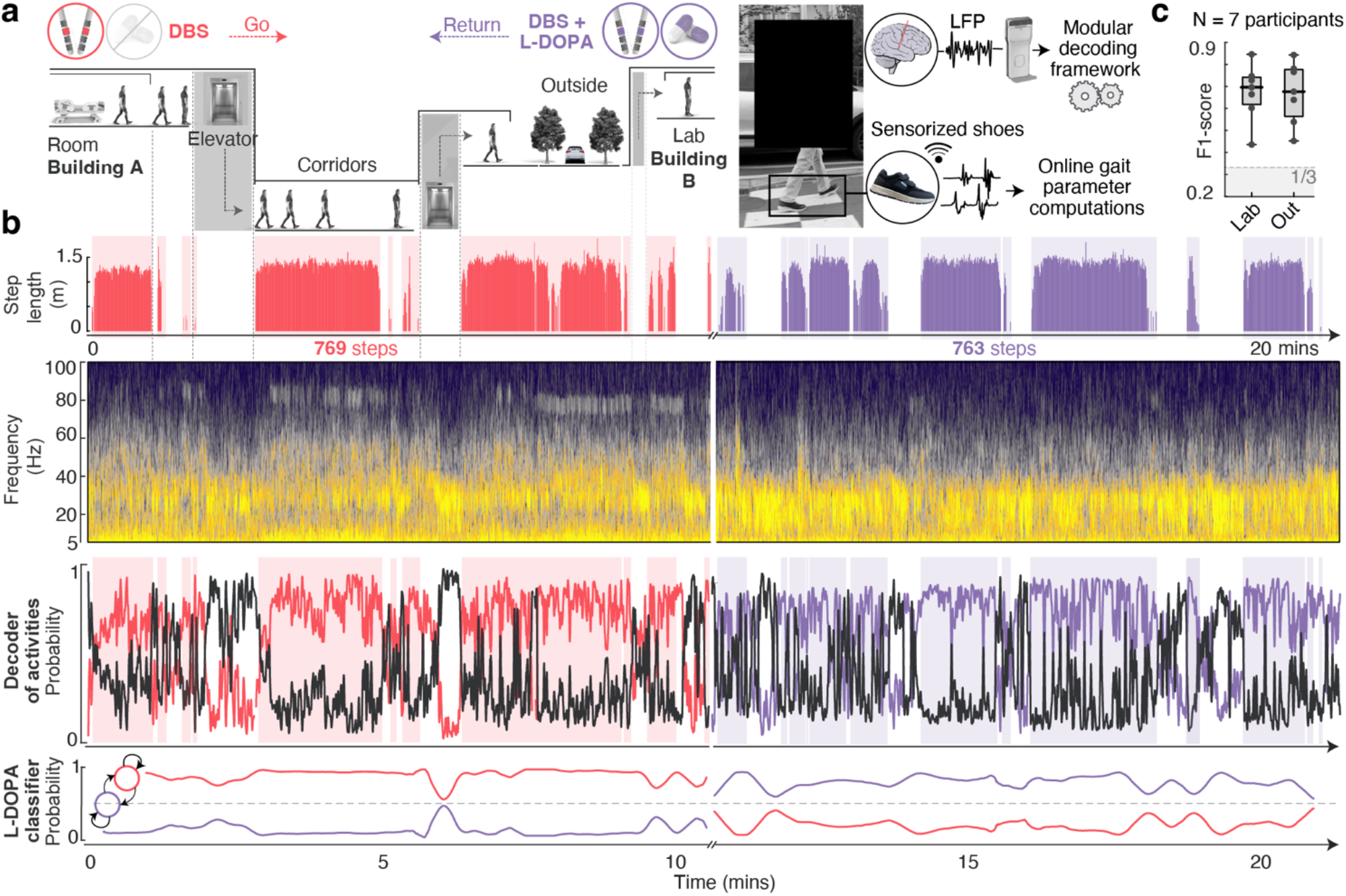
The modular decoding framework maintains prediction accuracy in out-of-laboratory settings. **(a)** Experimental setup to evaluate the real-world transferability of the modular decoding framework. Participants ambulated within hospital environments from building A to building B (~1km), navigating crowded hallways, elevators, and outdoor environments. Locomotion was continuously tracked using sensorized shoes with embedded machine learning algorithms that detect walking events and compute gait parameters in real time, synchronized with STN LFP recordings. The outbound path was performed under STN DBS alone (medication withdrawn >12 h), and the return under the standard-of-care combination of STN DBS and L-DOPA (~90 minutes post-intake). **(b)** Illustrative example of decoding performance across real-world mobility (participant PD135). Top: Step length measured across >1,500 gait cycles during unconstrained walking. Middle: Corresponding raw STN LFP spectrogram Bottom: Decoder probabilities from the selected activity decoder, aligned to probabilities from the L-DOPA classifier over time. **(c)** Boxplot represents the performance (F1-score) of activity decoders trained under controlled laboratory conditions and tested during real-world ambulation, across the 7 participants recorded.

**Extended Data Figure 9.**
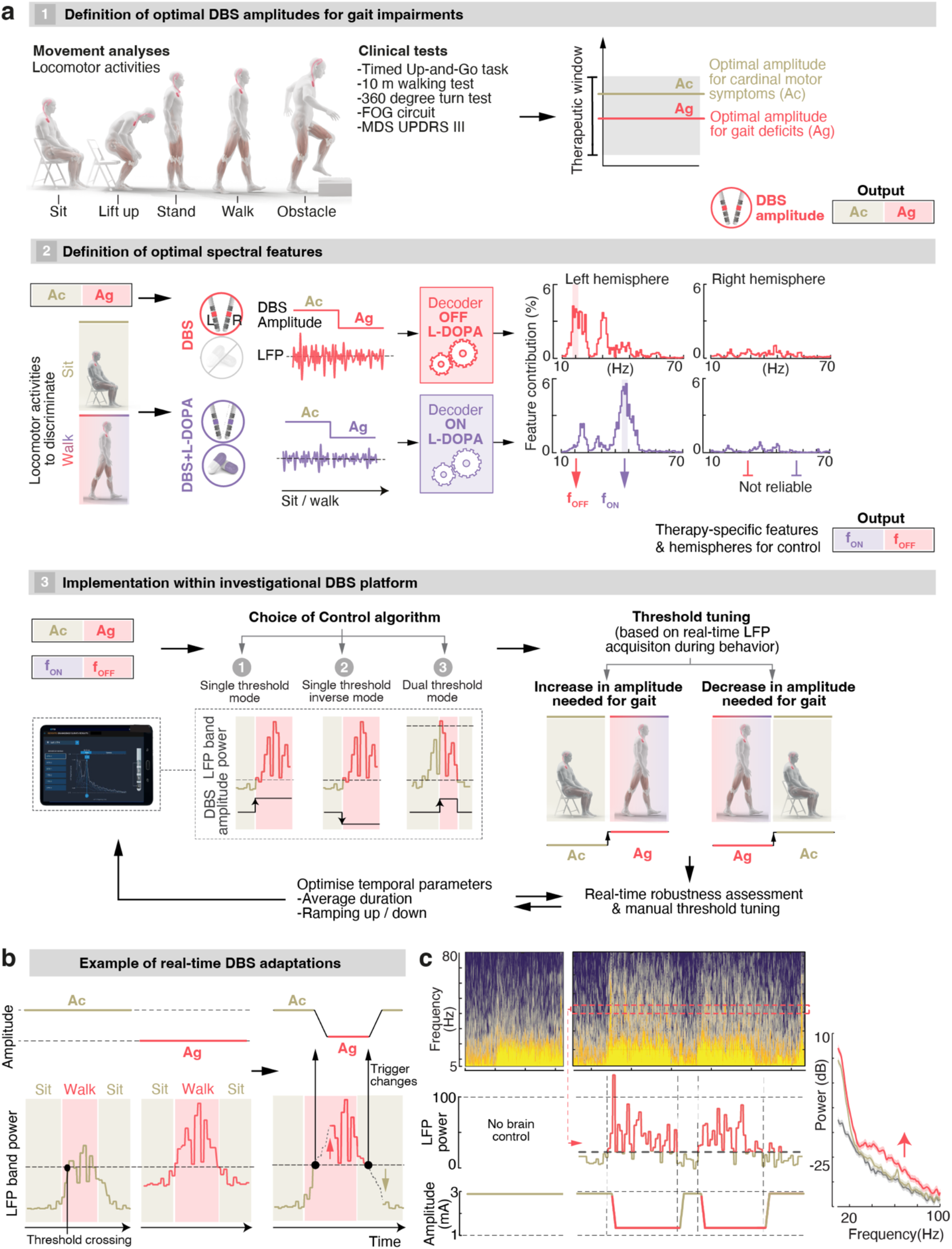
Implementation workflow for activity-dependent adaptive DBS. **(a)** The workflow is organized into three steps. First, we identified patient-specific locomotor deficits under the standard-of-care DBS amplitude and determined the optimal DBS amplitude to address these deficits. The impact of the new DBS amplitude on cardinal motor symptoms was also evaluated. Second, we identified the optimal spectral features for discriminating locomotor activities. Thes features were computed for each hemisphere, separately under high and low L-DOPA conditions, but across varying DBS amplitudes to ensure stability during real-time modulation of stimulation amplitude. Third, we implemented the selected parameters into the investigational DBS platform. This involved selecting the control mode that enabled up- or down-regulation of stimulation amplitude based on the chosen neural biomarkers, and tuning the triggering threshold and temporal parameters to maximize robustness. **(b)** Scheme illustrating how threshold tuning must capture movement-related modulations in STN LFPs across both therapeutic conditions (under standard-of-care DBS amplitude and at the optimal amplitude required to address gait deficits), and how real-time DBS adaptations based on this threshold influence band power. **(c)** Representative example of changes in STN dynamics during movement (participant P3), shown under both standard-of-care therapy and activity-dependent DBS therapy. Activity-dependent adaptations in DBS amplitude are triggered using gamma power (40Hz) as control signal.

**Extended Data Figure 10.**
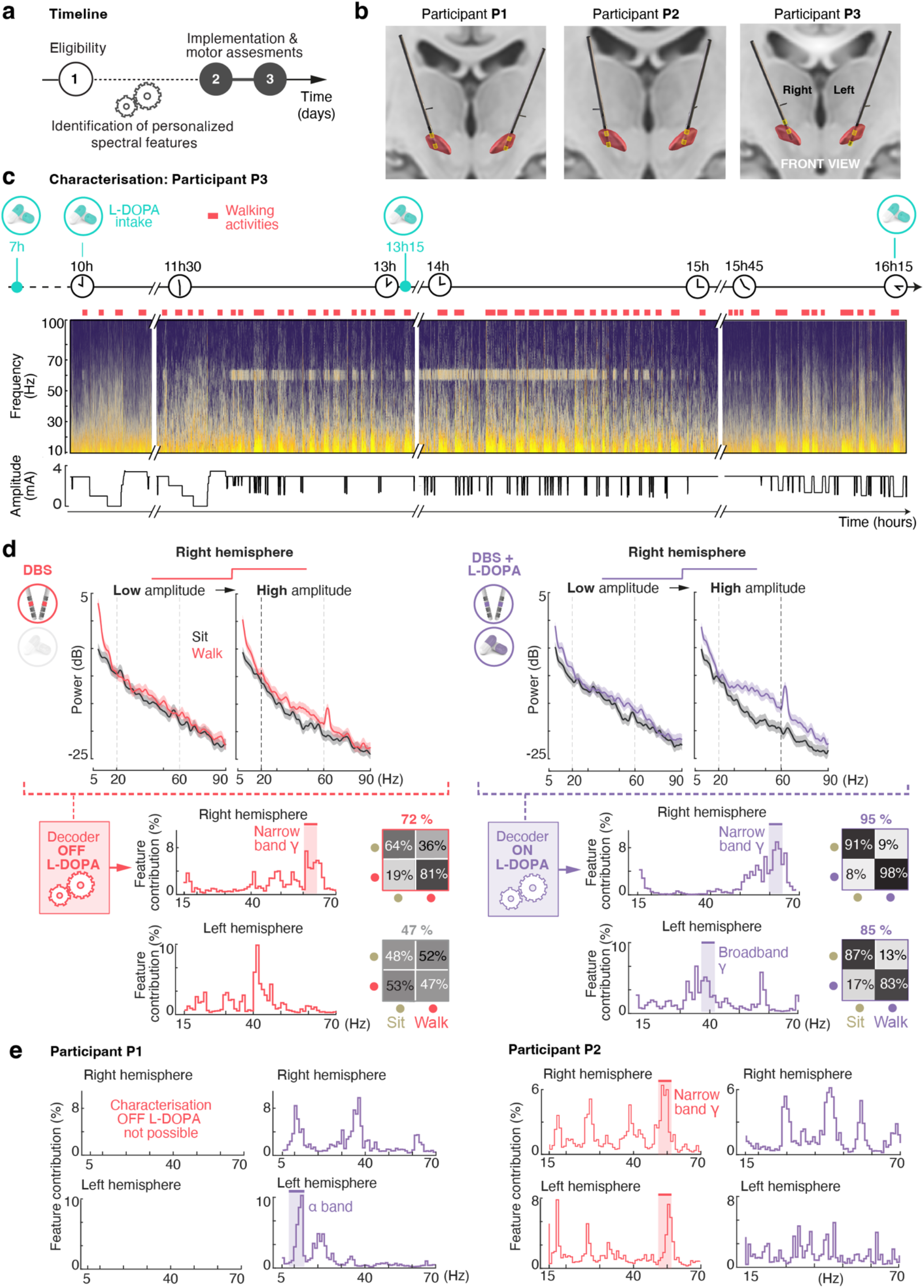
Patient-specific tuning of activity-dependent adaptive DBS based on individualized physiological decoding. **(a)** Timeline of the feasibility study. During the eligibility session, we assessed each participant’s cardinal motor symptoms, locomotor deficits, impedance values, and the quality of STN LFP signals during a battery of gait tasks. The modular decoding framework was then applied to extract individualized neural signatures of locomotor states across varying stimulation amplitudes and dopaminergic states. Participants meeting the inclusion criteria proceeded to two additional sessions to evaluate the safety and preliminary efficacy of activity-dependent adaptive DBS. **(b)** Anatomical reconstruction of electrode placement and recording contacts used for LFP acquisition in each participant included in the study. **(c)** Representative example of STN LFP recordings acquired across a full session, acquired across evolving L-DOPA conditions and varying STN DBS amplitudes, both at rest and during walking tasks. **(d)** Example of power spectral density (PSD) traces illustrating activity-dependent modulations in STN dynamics across L-DOPA conditions, for the two DBS amplitudes defined to target cardinal motor symptoms and locomotor deficits (participant P3, same as in panel **c**). In each condition, the modular decoder identified the most relevant spectral features for discriminating between locomotor states within the 7–70 Hz range, as permitted by the constraints of the DBS platform for real-time control. The lower bound was increased to 15 Hz in cases where low-frequency components raised concerns about walking-related artifacts. The optimal frequency band for each hemisphere (highlighted in shaded red or purple) was selected from among those suggested by the decoders, evaluated during online assessments, and iteratively refined based on real-time performance. When spectral features were similar across low- and high-L-DOPA conditions, the same frequency band was selected to enhance the stability of real-time adaptations. In cases where decoding performance from one hemisphere was not sufficient (as derived from confusion matrices), a unilateral control signal was used to modulate DBS amplitudes bilaterally (see **Extended Data Table 2**). **(e)** Representation of spectral features for participants P1 and P2. Participant P1 could not be assessed in the low L-DOPA condition due to motor impairment preventing completion of the necessary clinical evaluations. P3 only exhibited severe gait deficits (FOG) when low in L-DOPA. The selected frequency bands corresponded to the most informative spectral features in that therapeutic condition.

**Extended Data Figure 11.**
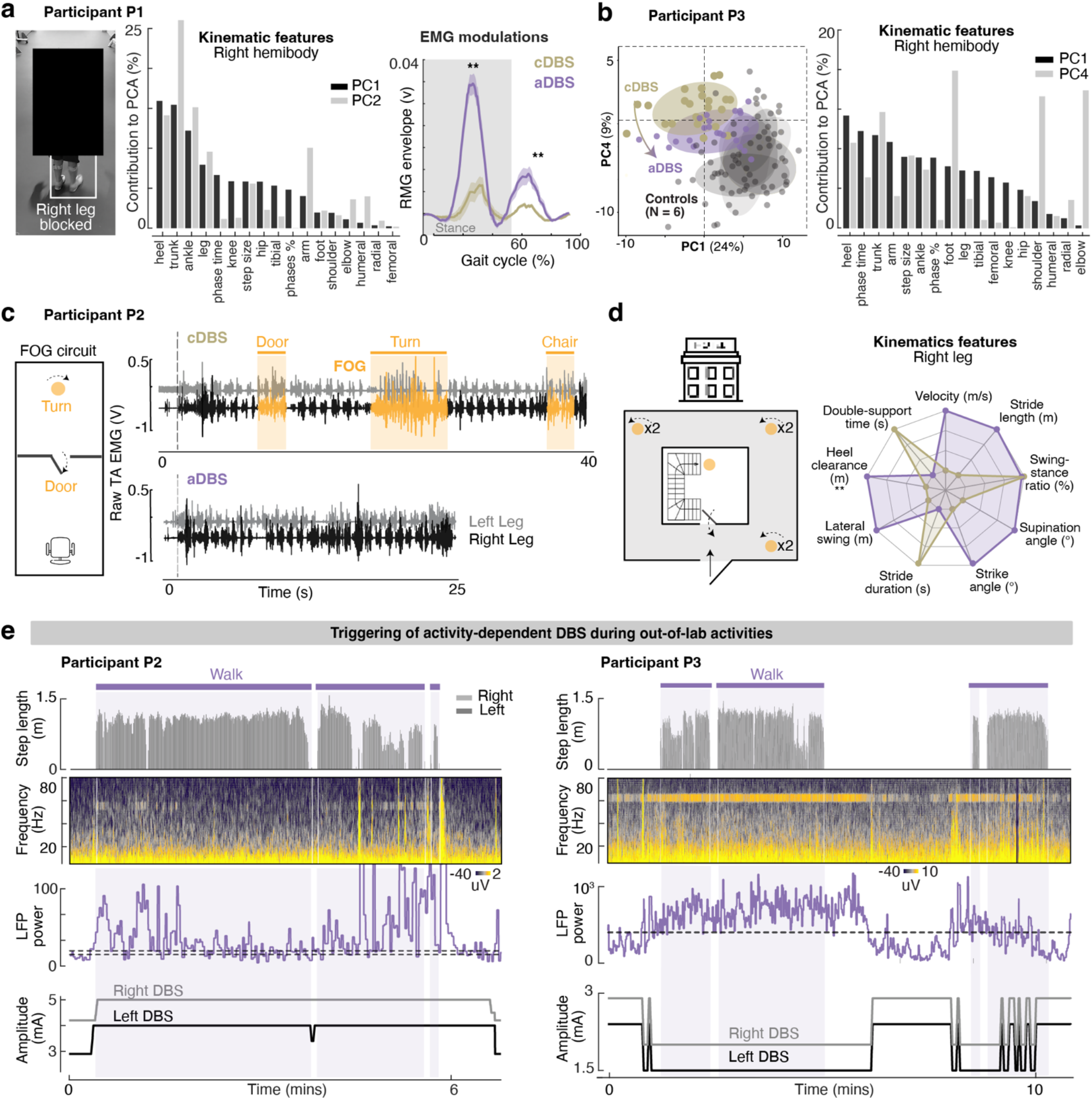
Participant-specific improvements in locomotor deficits mediated by activity-dependent DBS. **(a)** Illustrative screenshots showing Participant P1 with impaired gait fluidity due to reduced right leg flexion. Key kinematic features, derived from video-based motion tracking and used for principal component analysis (as shown in **Fig. 6d**), are grouped into biomechanical ‘domains’ (e.g., heel, ankle, knee, and hip joint, timing and proportion of gait phases). The percentage contribution of each domain to the first two principal components confirms predominant involvement of lower-leg and trunk parameters. EMG envelope traces (median ± SEM) from the right medial gastrocnemius (RMG) confirm increased muscle activation in the impaired limb. **(b)** PCA-based kinematic analysis during walking confirms gait improvements in Participant P3. The two principal components that best illustrate changes with aDBS with respect to standard-of-care DBS are shown. Under activity-dependent adaptive DBS (aDBS), gait patterns shift closer to those of healthy controls, indicating improved gait quality and reduction of disabling right foot and leg dyskinesia. The kinematic features that contribute the most to PC1 include heel kinematics and temporal gait parameters, reflecting enhanced rhythm and stability. Instead, PC4 primarily captured foot, shoulder, and elbow kinematics, likely reflecting compensatory upper-limb movements associated with foot dyskinesia. **(c)** Customized FOG circuit involving sit-to-stand transitions, doorways, and turns to elicit freezing of gait (FOG) in participant P2. Representative raw EMG traces of the tibialis anterior (TA) are shown for one repetition of the FOG circuit performed under standard-of-care DBS and one under activity-dependent adaptive DBS. The circuit was repeated twice per condition. The duration of FOG episodes was quantified from bilateral EMG activity patterns (FOG events summarized as individual trials in **Fig. 6e)**. The time to complete each freezing trial was computed as the interval between the final pre-event toe-off and the first post-event toe-off **(d)** Out-of-laboratory gait assessments in participant P2 were conducted along a predefined walking path within the hotel where the participant stayed between experimental sessions 2 and 3. The circuit included seven turning points (indicated by orange dots), a door crossing, and a staircase descent. The proportion of turns affected by FOG and the time required to complete each turn are quantified in **Fig. 6i**. Kinematic gait features extracted from the sensorized shoes (NUSHU, Magnes AG, Switzerland) further supported improvements in gait quality under activity-dependent DBS. **(e)** Illustrative recordings of sustained activity-dependent DBS during out-of-laboratory assessments in participants P2 and P3. Panels display aligned step length trajectories (top), raw STN LFP spectrograms (5–80 Hz, middle), LFP band-power used to trigger adaptations in DBS amplitude (narrow-band gamma: 55.6 Hz for P2; 62.5 Hz for P3), and corresponding stimulation adjustments (bottom). DBS amplitude was increased in P2 to mitigate locomotor deficits, whereas it was decreased in P3. As such, the dual-threshold mode was used to control DBS adaptations in P2, whereas the single-threshold inverse mode was employed in P3. The temporal parameters (average duration, blanking period and onset duration) controlling the responsiveness of aDBS algorithms were tuned to compensate for fluctuations in the stability of the control band, which varied with task complexity and physical effort (e.g., outdoors walking up or down, stairs etc). In addition, the dual threshold in P2 allowed to further compensate for variability in the control band. Unpaired t-tests were performed to compare aDBS and cDBS conditions (*p < 0.05, **p < 0.01), with Bonferroni correction applied for multiple comparisons.

**Extended Data Table 1.**
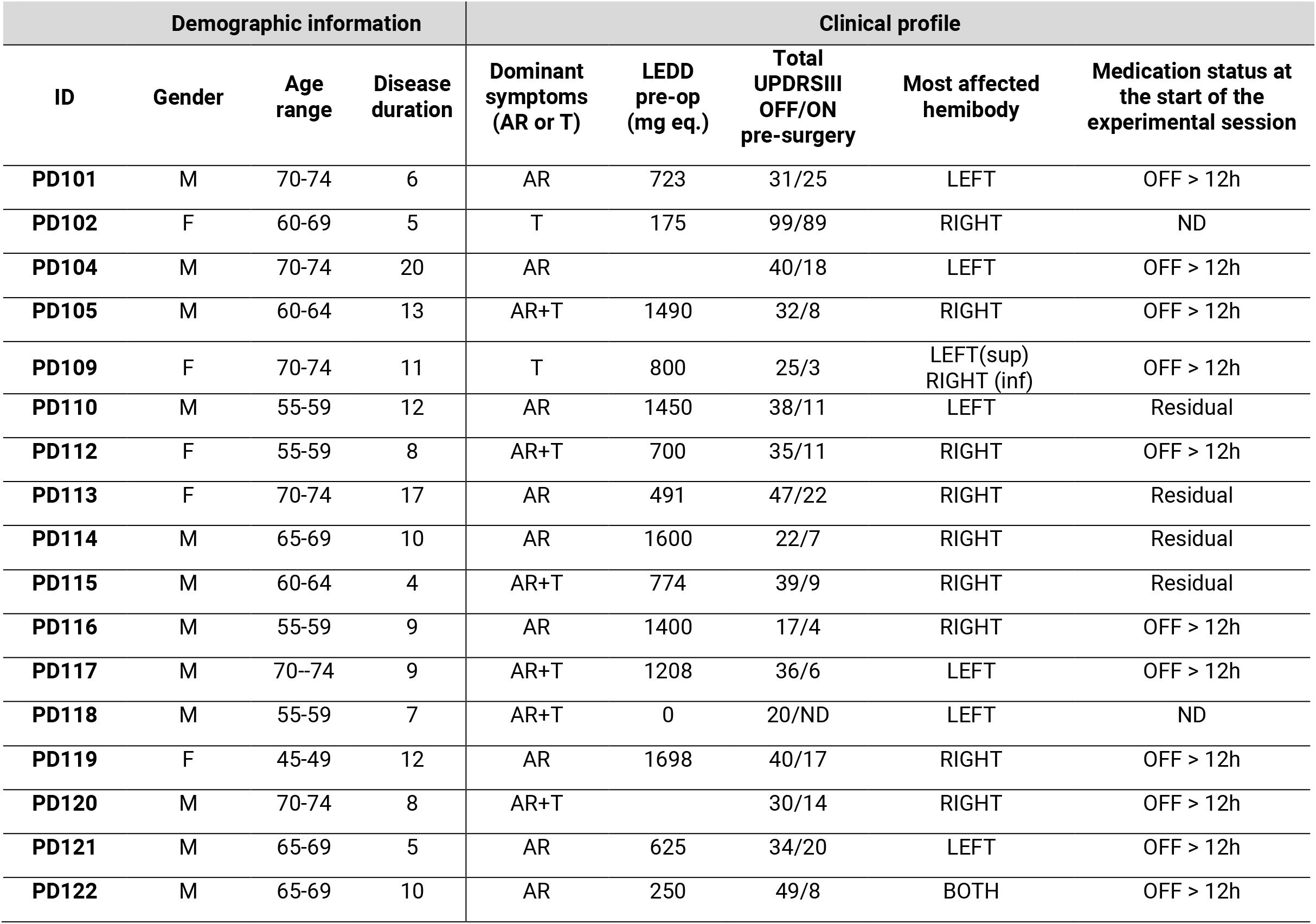
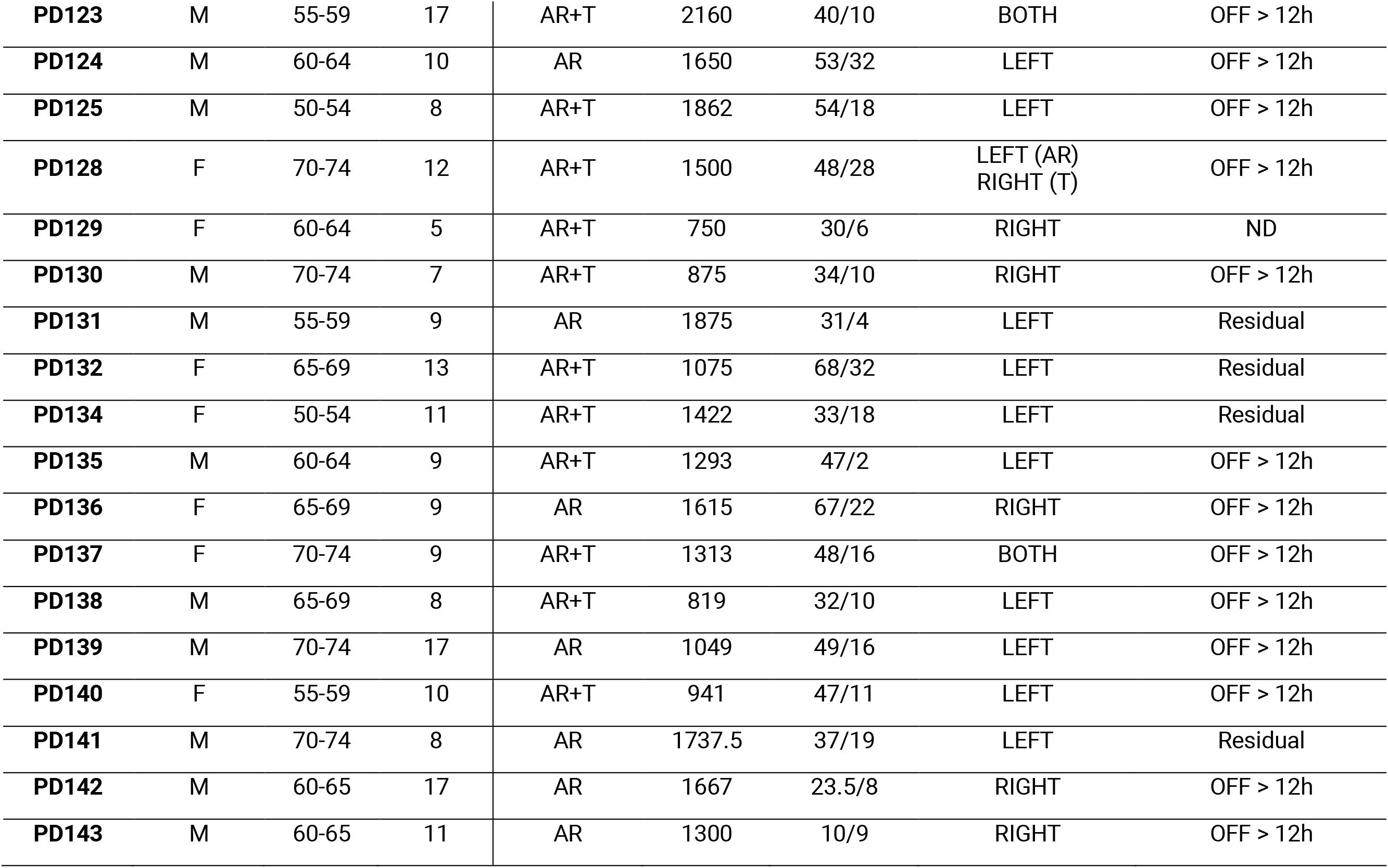
Demographic and clinical characteristics of all 35 participants included in the characterization of physiological principles underlying therapy-specific encoding of locomotor activities in STN dynamics, and the development of neural decoders. (AR: Akinetic-rigid, T: Tremor, ND: No Dopamine)

**Extended Data Table 2.**
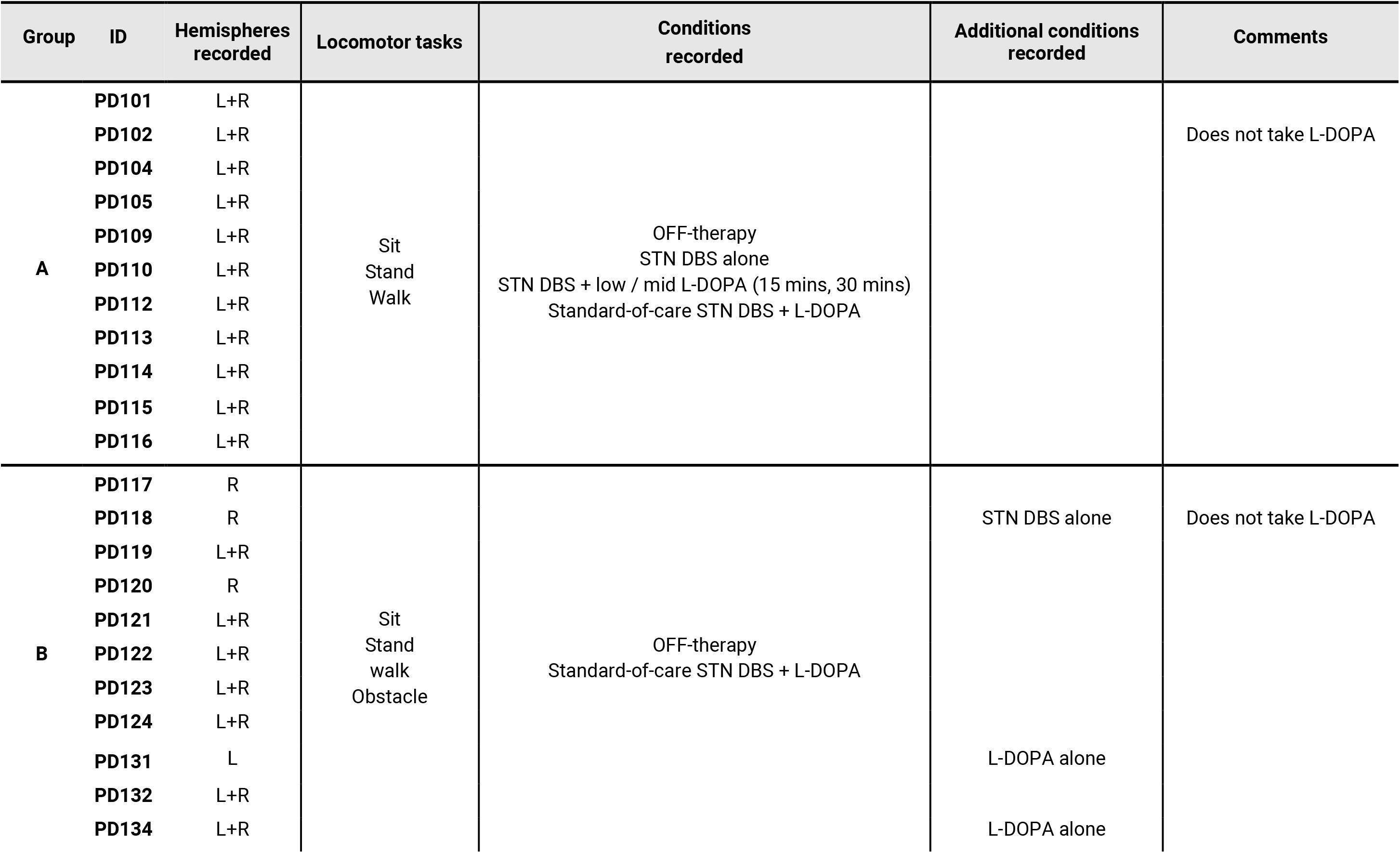
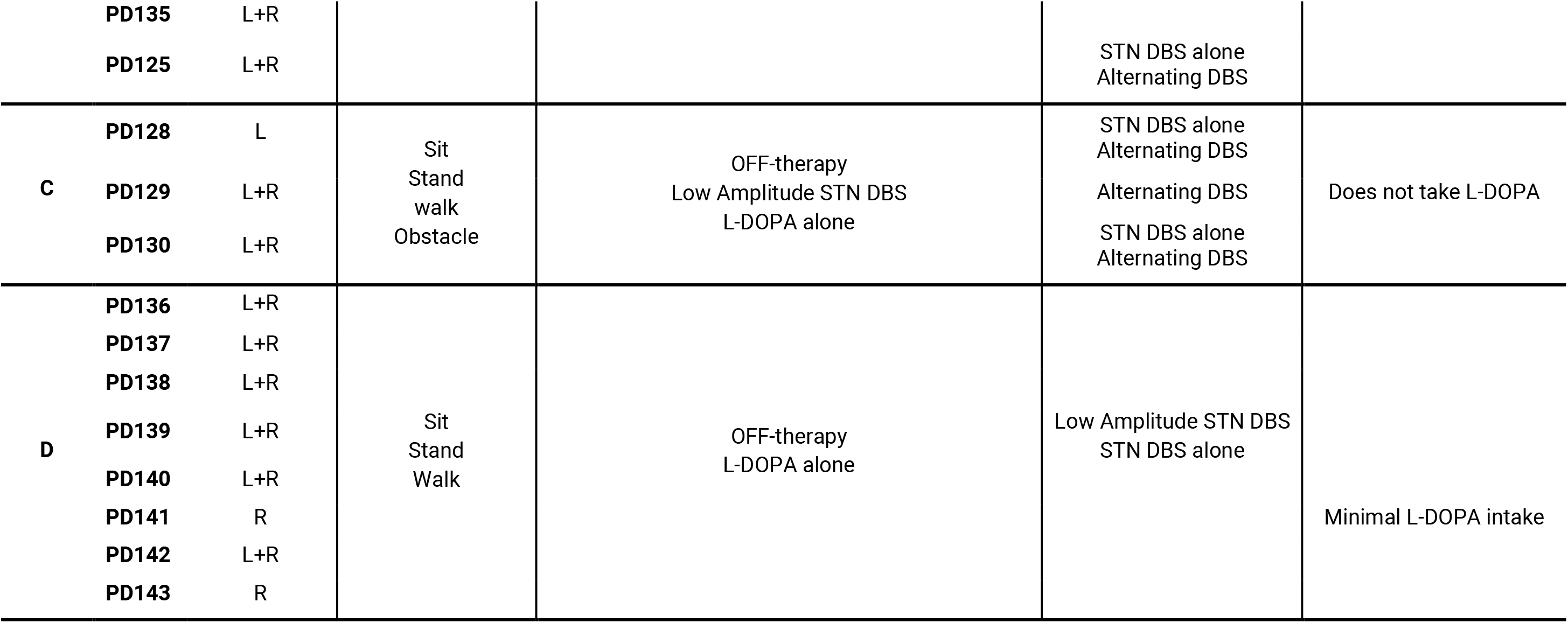
Locomotor tasks and therapeutic conditions recorded in all 35 participants included in the characterization of the physiological principles underlying therapy-specific encoding of locomotor activities in STN dynamics, and leveraged for the development of neural decoders.

**Extended Data Table 3.** Demographic and clinical characteristics of the 7 participants recruited for the proof-of-concept feasibility study assessing the safety and preliminary efficacy of activity-dependent adaptive DBS. This includes MDS-UPDRS Part III scores (Movement Disorder Society Unified Parkinson’s Disease Rating Scale) assessed in the low dopaminergic state. Participants P1, P2 and P3 met the eligibility criteria after Session 1 and were enrolled in the interventional phase of the study. For these participants, the most bothersome gait deficits targeted with activity-dependent adaptive DBS are reported, along with the affected body side and the corresponding Freezing of Gait Questionnaire (FOG-Q) score (range: 0–24), when FOG was among the presenting symptoms.

| Demographic information |  |  |  |  |  |  |  | Gait deficit |  |  |  |
| --- | --- | --- | --- | --- | --- | --- | --- | --- | --- | --- | --- |
| Participant | Age range | Gender | Disease duration (yrs) | Months from DBS implant (range) | Type | MDS-UPDRS III Low MED | Eligibility matched | First | Second | Body side | FOGQ |
| P1 | 60-64 | F | 13 | 5-9 | Akinetic-rigid and tremor | 41 | YES | Blocked leg | — | Right | — |
| P2 | 60-64 | M | 11 | 15-19 | Akinetic-rigid | 29 | YES | FoG | — | Right (sup)<br>Left (inf) | 12 |
| P3 | 60-64 | M | 12 | 10-14 | Akinetic-rigid | 14 | YES | FoG | Lower limb dyskinesia | Right | 12 |
| P4 | 50-54 | F | 13 | 0-4 | Akinetic-rigid and tremor | 25 | NO | — | — | — | — |
| P5 | 65-59 | M | 18 | 0-4 | Akinetic-rigid and tremor | 40 | NO | — | — | — | — |
| P6 | 70-74 | M | 12 | 30-34 | Akinetic-rigid and tremor | 37 | NO | — | — | — | — |
| P7 | 60-64 | M | 16 | 40-44 | Akinetic-rigid | 26 | NO | — | — | — | — |

**Extended Data Table 4.**
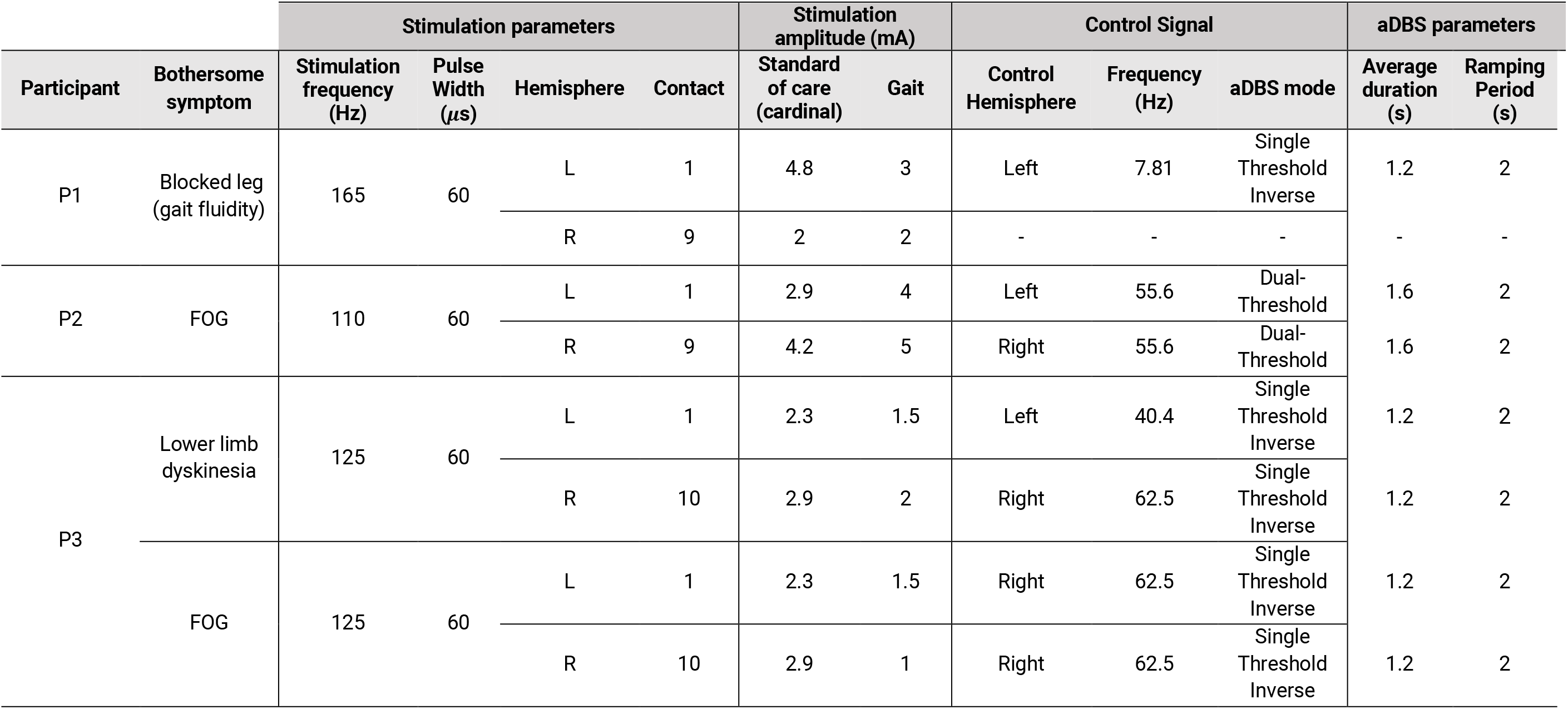
STN DBS parameters used for activity-dependent adaptive DBS targeting locomotor deficits, compared to standard-of-care continuous DBS. All participants were stimulated using monopolar configurations. The monitored frequency corresponds to the center of the 5-Hz band selected during participant-specific biomarker identification. In participant P1, activity-dependent adaptive DBS was delivered unilaterally in the left hemisphere to target the more affected right hemibody. In participant P3, bilateral stimulation was controlled via sensing from a single hemisphere, with sensing disabled contralaterally to allow this configuration.

| Participant | Bothersome symptom | Stimulation parameters |  |  |  | Stimulation amplitude (mA) |  | Control Signal |  |  | aDBS parameters |  |
| --- | --- | --- | --- | --- | --- | --- | --- | --- | --- | --- | --- | --- |
| | | Stimulation frequency (Hz) | Pulse Width (μs) | Hemisphere | Contact | Standard of care (cardinal) | Gait | Control Hemisphere | Frequency (Hz) | aDBS mode | Average duration (s) | Ramping Period (s) |
| P1 | Blocked leg (gait fluidity) | 165 | 60 | L | 1 | 4.8 | 3 | Left | 7.81 | Single Threshold Inverse | 1.2 | 2 |
|  |  |  |  | R | 9 | 2 | 2 | - | - | - | - | - |
| P2 | FOG | 110 | 60 | L | 1 | 2.9 | 4 | Left | 55.6 | Dual-Threshold | 1.6 | 2 |
|  |  |  |  | R | 9 | 4.2 | 5 | Right | 55.6 | Dual-Threshold | 1.6 | 2 |
| P3 | Lower limb dyskinesia | 125 | 60 | L | 1 | 2.3 | 1.5 | Left | 40.4 | Single Threshold Inverse | 1.2 | 2 |
|  |  |  |  | R | 10 | 2.9 | 2 | Right | 62.5 | Single Threshold Inverse | 1.2 | 2 |
|  | FOG | 125 | 60 | L | 1 | 2.3 | 1.5 | Right | 62.5 | Single Threshold Inverse | 1.2 | 2 |
|  |  |  |  | R | 10 | 2.9 | 1 | Right | 62.5 | Single Threshold Inverse | 1.2 | 2 |

